# Pathophysiology-based subphenotyping of individuals at elevated risk for type 2 diabetes

**DOI:** 10.1101/2020.10.12.20210062

**Authors:** Robert Wagner, Martin Heni, Adam G. Tabak, Jürgen Machann, Fritz Schick, Elko Randrianarisoa, Martin Hrabě de Angelis, Andreas L. Birkenfeld, Norbert Stefan, Andreas Peter, Hans-Ulrich Häring, Andreas Fritsche

## Abstract

The state of intermediate hyperglycemia is indicative of elevated risk of developing type 2 diabetes^1^. However, the current definition of prediabetes neither reflects subphenotypes of pathophysiology of type 2 diabetes nor is it predictive of future metabolic trajectories. We used partitioning on variables derived from oral glucose tolerance tests, MRI measured body fat distribution, liver fat content, and genetic risk in a cohort of extensively phenotyped individuals who are at increased risk for type 2 diabetes^2,3^ to identify six distinct clusters of subphenotypes. Three of the identified subphenotypes have increased glycemia (clusters 3, 5 and 6), but only individuals in clusters 5 and 3 have immanent diabetes risks. By contrast, those in cluster 6 have moderate risk of type 2 diabetes, but an increased risk of kidney disease and all-cause mortality. Findings were replicated in an independent cohort using simple anthropomorphic and glycemic constructs^4^. This proof-of-concept study demonstrates that pathophysiological heterogeneity exists before diagnosis of type 2 diabetes and highlights a group of individuals who have an increased risk of complications without rapid progression to overt type 2 diabetes.

## Introduction

Type 2 diabetes occurs when insulin secretion from pancreatic beta-cells cannot sufficiently be increased to compensate for insulin resistance. Causes of beta-cell dysfunction and insulin resistance are heterogeneous, as are individual trajectories of hyperglycemia and subsequent manifestation of diabetes complications^5^. The currently used binary definition of type 2 diabetes is based solely on blood glucose and cannot differentiate between patients with mild or more aggressive disease, the latter of which is prone to early development of complications. In addition to blood glucose, new proposed diabetes classifications^6,7^ introduced additional variables, such as insulin secretion and insulin sensitivity, to sub-classify the type 2 diabetes spectrum with the primary aim of a better prediction of metabolic dysfunction and complications. The development of type 2 diabetes is a slow process, and its manifestation is preceded by a phase of prediabetes which often remains undiagnosed. Some diabetes complications, such as the unexpectedly frequent early diabetic kidney disease in the newly identified severe insulin resistant diabetes cluster^6^, might require preventive actions prior to the clinical manifestation of type 2 diabetes. The assessment of insulin secretion and insulin sensitivity could be hindered by secondary gluco-lipotoxicity, once diabetes has developed and glucose levels are continuously elevated^8^. Determination of prediabetes subphenotypes prior to the manifestation of diabetes could improve detection of individuals at risk for diabetes and complications. Using accurate measurements of insulin sensitivity and insulin secretion based on oral glucose tolerance test (OGTT)-derived variables, as well as variables linked to diabetes pathogenesis, we describe a novel subphenotyping approach of metabolic risk before diabetes manifestation. Variables include HDL-cholesterol, which has been causally linked to type 2 diabetes^9^, MR-imaging-derived measures of metabolically unfavorable and favorable fat compartments^10^ and liver fat content measured with ^1^H-MR-spectroscopy. To assess genetic liability, we also incorporated a type 2 diabetes polygenic risk score^11^ as partitioning variable. The clusters identified by the sophisticated phenotypes in the TUEF/TULIP cohort were replicated using simpler markers of similar anthropometric and glycemic constructs in a large prospective occupational cohort (the Whitehall II study)^4^. Our results suggest that stratification of populations at increased risk for type 2 diabetes using simple clinical features could allow for precise and efficient prevention strategies individuals at increased risk of developing type 2 diabetes.

## Results

Initial clustering and identification of the subphenotypes was done using data from a subset of participants (n=899) from the Tuebingen Family study and Tuebingen Lifestyle Program (TUEF/TULIP) study. Analysis was performed on data for participants who had no missing values for the preselected phenotyping variables: glucose challenge; insulin sensitivity; insulin secretion; HDL-cholesterol; liver fat content; subcutaneous fat volume; visceral fat volume; and a polygenic risk score for type 2 diabetes risk The clustering was replicated in the Whitehall II cohort (n=6810) using conceptually similar variables: glycemia during glucose challenge, insulin sensitivity, insulin secretion, fasting insulin, fasting triglycerides, waist circumference, hip circumference, BMI and HDL-cholesterol (Extended Data 1; see Methods). We identified six clusters with distinctive patterns of the variables in the TUEF/TULIP study (Figure 1.A,B), which were replicated in the Whitehall II cohort (Figure 1 C,D). Cluster characteristics and comparisons are shown in Table 1, Suppl.Table 1-3 and key features of the clusters are reported in Extended Data 2.

**Table 1.** Cluster characteristics of the TUEF/TULIP cohort after stratification for the 6 clusters. P-values were computed with one-way ANOVA for continuous variables and two-sided chi-squared tests for categorical variables.

|  | 1 | 2 | 3 | 4 | 5 | 6 | p-value |
| --- | --- | --- | --- | --- | --- | --- | --- |
|  | Low risk | Very low risk | Beta-cell failure | Low risk obese | High risk insulin resistant fatty liver | High risk visceral fat nephropathy |  |
| **n** | 173 | 154 | 146 | 153 | 91 | 182 |  |
| **sex = male (%)** | 64 (37.0) | 59 (38.3) | 65 (44.5) | 56 (36.6) | 35 ( 38.5) | 67 (36.8) | 0.72 |
| **age (mean (SD))** | 39.05<br>(12.55) | 41.75<br>(13.29) | 52.26<br>(12.11) | 40.14<br>(11.85) | 49.74<br>(11.81) | 47.38 (12.64) | $8.7 \times 10^{-27}$ |
| **BMI (kg/m<sup>2</sup>) (mean (SD))** | 26.82<br>(3.16) | 23.45<br>(3.32) | 29.15<br>(4.01) | 31.54<br>(3.67) | 34.45<br>(5.11) | 34.94 (4.90) | $1.6 \times 10^{-135}$ |
| **waist circumference (cm) (mean (SD))** | 88.44<br>(9.63) | 80.58<br>(9.80) | 97.11<br>(11.21) | 99.14<br>(10.59) | 108.17<br>(12.88) | 107.86 (12.34) | $1 \times 10^{-111}$ |
| **hip circumference (cm) (mean (SD))** | 101.62<br>(7.71) | 95.66<br>(8.01) | 105.80<br>(13.25) | 112.61<br>(9.01) | 115.17<br>(11.02) | 117.06 (10.58) | $3.1 \times 10^{-89}$ |
| **total adipose tissue MRI (liter) (mean (SD))** | 27.71<br>(6.42) | 20.75<br>(7.63) | 33.27<br>(9.98) | 42.34<br>(9.31) | 46.20<br>(12.07) | 48.28 (12.00) | $4.4 \times 10^{-142}$ |
| **sq adipose tissue MRI (liter) (mean (SD))** | 8.78 (3.13) | 5.96 (3.50) | 10.95<br>(4.33) | 15.02<br>(4.79) | 16.72<br>(5.75) | 18.13 (6.19) | $1.6 \times 10^{-111}$ |
| **visceral adipose tissue MRI (liter) (mean (SD))** | 2.40 (1.48) | 1.77 (1.21) | 4.16<br>(1.92) | 3.75 (1.97) | 5.73<br>(2.34) | 5.64 (2.44) | $1.9 \times 10^{-87}$ |
| **sq to visceral adipose ratio (mean (SD))** | 5.16 (3.13) | 4.63 (2.84) | 3.38<br>(2.18) | 5.38 (3.18) | 3.33<br>(1.56) | 3.78 (1.95) | $5 \times 10^{-15}$ |
| **visceral adipose % of total (mean (SD))** | 0.09 (0.06) | 0.09 (0.06) | 0.13<br>(0.06) | 0.09 (0.06) | 0.13<br>(0.05) | 0.12 (0.06) | $5.9 \times 10^{-17}$ |
| **liver fat content (mean (SD))** | 3.34 (3.25) | 2.16 (2.90) | 5.10<br>(3.72) | 3.61 (3.51) | 20.79<br>(5.73) | 9.88 (5.49) | $5.2 \times 10^{-193}$ |
| **fatty-liver disease (%) = yes (%)** | 28 (16.2) | 8 ( 5.2) | 51 (34.9) | 26 (17.0) | 91<br>(100.0) | 137 (75.3) | $3.1 \times 10^{-82}$ |
| **renal sinus fat (mean of r&l, %**<br>**(mean (SD))** | 5.20 (3.80) | 5.77 (4.23) | 9.42<br>(4.75) | 7.15 (4.52) | 10.02<br>(4.87) | 12.07 (6.08) | 1.3×10<sup>-26</sup> |
| **systolic blood pressure (mmHg)**<br>**(mean (SD))** | 126.26<br>(14.15) | 123.36<br>(15.81) | 135.73<br>(18.59) | 126.32<br>(15.43) | 143.66<br>(19.34) | 137.86 (17.06) | 6.3×10<sup>-29</sup> |
| **diastolic blood pressure (mmHg)**<br>**(mean (SD))** | 80.25<br>(10.64) | 78.52<br>(11.09) | 84.93<br>(12.07) | 81.16<br>(10.14) | 92.57<br>(13.62) | 87.47 (12.07) | 2.5×10<sup>-24</sup> |
| **heart rate (mean (SD))** | 69.35<br>(10.00) | 67.47<br>(10.85) | 69.29<br>(9.91) | 68.13<br>(10.76) | 75.39<br>(12.26) | 72.26 (9.64) | 1.9×10<sup>-08</sup> |
| **fasting glucose (mmol/l) (mean**<br>**(SD))** | 5.12 (0.44) | 5.04 (0.50) | 5.64<br>(0.55) | 5.14 (0.41) | 5.93<br>(0.58) | 5.48 (0.50) | 1.6×10<sup>-56</sup> |
| **post-challenge glucose (mmol/l)**<br>**(mean (SD))** | 6.12 (1.09) | 5.99 (1.26) | 7.87<br>(1.38) | 5.72 (0.86) | 8.31<br>(1.54) | 7.10 (1.38) | 1.2×10<sup>-80</sup> |
| **glycaemic category (%)** |  |  |  |  |  |  | 2.3×10<sup>-68</sup> |
| **NGT** | 139 (80.3) | 126 (81.8) | 36 (24.7) | 131 (85.6) | 12 ( 13.2) | 85 (46.7) |  |
| **IFG** | 24 (13.9) | 17 (11.0) | 41 (28.1) | 20 (13.1) | 20 (22.0) | 46 (25.3) |  |
| **IGT** | 8 (4.6) | 10 (6.5) | 36 (24.7) | 1 (0.7) | 14 (15.4) | 29 (15.9) |  |
| **IFG+IGT** | 2 (1.2) | 1 (0.6) | 33 (22.6) | 1 (0.7) | 45 (49.5) | 22 (12.1) |  |
| **GAD antibody = TRUE (%)** | 5 (3.2) | 4 (2.8) | 3 (2.5) | 5 (3.7) | 2 (2.6) | 9 (5.7) | 0.7 |
| **glycated haemoglobin (mmol/mol)**<br>**(mean (SD))** | 35.67<br>(4.47) | 36.77<br>(4.03) | 38.95<br>(6.37) | 35.80<br>(3.95) | 40.06<br>(3.60) | 38.23 (3.86) | 3.1×10<sup>-19</sup> |
| **triglycerides (mmol/l) (mean (SD))** | 1.26 (0.57) | 0.87 (0.35) | 1.59<br>(1.17) | 1.16 (0.63) | 2.04<br>(1.13) | 1.57 (0.79) | 2.3×10<sup>-30</sup> |
| **insulin sensitivity (Matsuda)**<br>**(mean (SD))** | 14.54<br>(6.07) | 24.33<br>(9.08) | 11.52<br>(5.39) | 17.63<br>(7.16) | 5.99<br>(3.01) | 7.46 (3.78) | 5.3×10<sup>-128</sup> |
| **fasting insulin (pmol/l) (mean**<br>**(SD))** | 51.97<br>(22.02) | 32.34<br>(14.35) | 54.89<br>(25.73) | 48.55<br>(22.18) | 113.98<br>(64.09) | 99.81 (48.55) | 5.5×10<sup>-93</sup> |
| **insulinogenic index (mean (SD))** | 184.24<br>(274.80) | 100.61<br>(139.56) | 69.84<br>(37.14) | 153.66<br>(136.01) | 125.06<br>(69.77) | 191.29<br>(136.68) | 1.9×10<sup>-13</sup> |
| **disposition index (mean (SD))** | 2804.28<br>(6133.07) | 2485.30<br>(5193.75) | 701.59<br>(293.53) | 2475.65<br>(2227.36) | 653.97<br>(357.25) | 1270.95<br>(979.49) | 1.5×10<sup>-09</sup> |
| **C-reactive protein (mg/dl) (mean**<br>**(SD))** | 0.20 (0.34) | 0.12 (0.25) | 0.21<br>(0.34) | 0.29 (0.32) | 0.49<br>(0.47) | 0.39 (0.42) | 2×10<sup>-18</sup> |
| **cholesterol (mmol/l) (mean (SD))** | 4.91 (0.95) | 4.88 (0.87) | 5.27<br>(1.02) | 4.82 (0.98) | 5.34<br>(0.93) | 5.14 (0.93) | 2.2×10<sup>-06</sup> |
| **LDL (mmol/l) (mean (SD))** | 3.04 (0.89) | 2.73 (0.78) | 3.22<br>(0.85) | 2.97 (0.82) | 3.41<br>(0.84) | 3.15 (0.80) | 2.1×10<sup>-09</sup> |
| **HDL (mmol/l) (mean (SD))** | 1.34 (0.28) | 1.69 (0.36) | 1.32<br>(0.29) | 1.27 (0.29) | 1.18<br>(0.27) | 1.28 (0.30) | $2.8 \times 10^{-45}$ |
| **aspartate-aminotransferase (U/l)**<br>**(mean (SD))** | 22.38<br>(6.98) | 22.79<br>(7.91) | 22.52<br>(7.03) | 22.14<br>(6.97) | 32.73<br>(14.50) | 25.18 (9.94) | $3.8 \times 10^{-21}$ |
| **alanine-aminotransferase (U/l)**<br>**(mean (SD))** | 24.95<br>(13.39) | 22.41<br>(10.12) | 25.42<br>(10.48) | 26.34<br>(15.06) | 48.47<br>(34.70) | 34.24 (18.43) | $3 \times 10^{-32}$ |
| **gamma-glutamyl transferase (U/l)**<br>**(mean (SD))** | 22.82<br>(19.52) | 18.24<br>(15.11) | 28.49<br>(26.08) | 21.48<br>(14.03) | 39.82<br>(34.49) | 33.90 (26.46) | $8 \times 10^{-16}$ |
| **serum creatinine (mg/dl) (mean**<br>**(SD))** | 0.83 (0.18) | 0.81 (0.17) | 0.82<br>(0.18) | 0.82 (0.15) | 0.78<br>(0.15) | 0.79 (0.17) | 0.18 |
| **urinary albumin-creatinine ratio**<br>**(mean (SD))** | 17.31<br>(35.16) | 18.46<br>(28.62) | 16.05<br>(14.97) | 17.58<br>(30.75) | 24.11<br>(45.77) | 16.51 (16.75) | 0.53 |
| **carotid intima media thickness**<br>**(mm) (mean (SD))** | 0.52 (0.12) | 0.53 (0.10) | 0.63<br>(0.13) | 0.54 (0.12) | 0.64<br>(0.12) | 0.60 (0.12) | $2.8 \times 10^{-13}$ |
| **polygenic risk score (mean (SD))** | -0.09<br>(0.97) | 0.15 (0.91) | 0.24<br>(0.92) | -0.17<br>(0.91) | 0.11<br>(0.81) | -0.07 (1.01) | 0.00057 |
| **family history of diabetes (%)** |  |  |  |  |  |  | 0.0084 |
| **a_no family history** | 64 (38.1) | 58 (39.5) | 42 (29.6) | 64 (42.7) | 27 (31.0) | 72 (41.1) |  |
| **b_second degree relative** | 37 (22.0) | 35 (23.8) | 22 (15.5) | 38 (25.3) | 15 (17.2) | 29 (16.6) |  |
| **c_first degree relative** | 67 (39.9) | 54 (36.7) | 78 (54.9) | 48 (32.0) | 45 (51.7) | 74 (42.3) |  |
| **ever smoked = yes (%)** | 86 (49.7) | 65 (42.2) | 82 (56.2) | 81 (52.9) | 45 (49.5) | 113 (62.1) | 0.011 |
| **current smoking = yes (%)** | 16 (9.9) | 8 (5.7) | 15 (11.0) | 12 (8.5) | 2 (2.4) | 18 (10.7) | 0.16 |
| **cholesterol lowering medication =**<br>**yes (%)** | 6 ( 3.5) | 0 ( 0.0) | 8 ( 5.5) | 2 ( 1.3) | 1 ( 1.1) | 6 ( 3.3) | 0.038 |
| **antihypertensive medication = yes**<br>**(%)** | 10 (5.8) | 3 (1.9) | 21 (14.4) | 2 (1.3) | 25 (27.5) | 44 (24.2) | $3.6 \times 10^{-17}$ |

**Figure 1.**
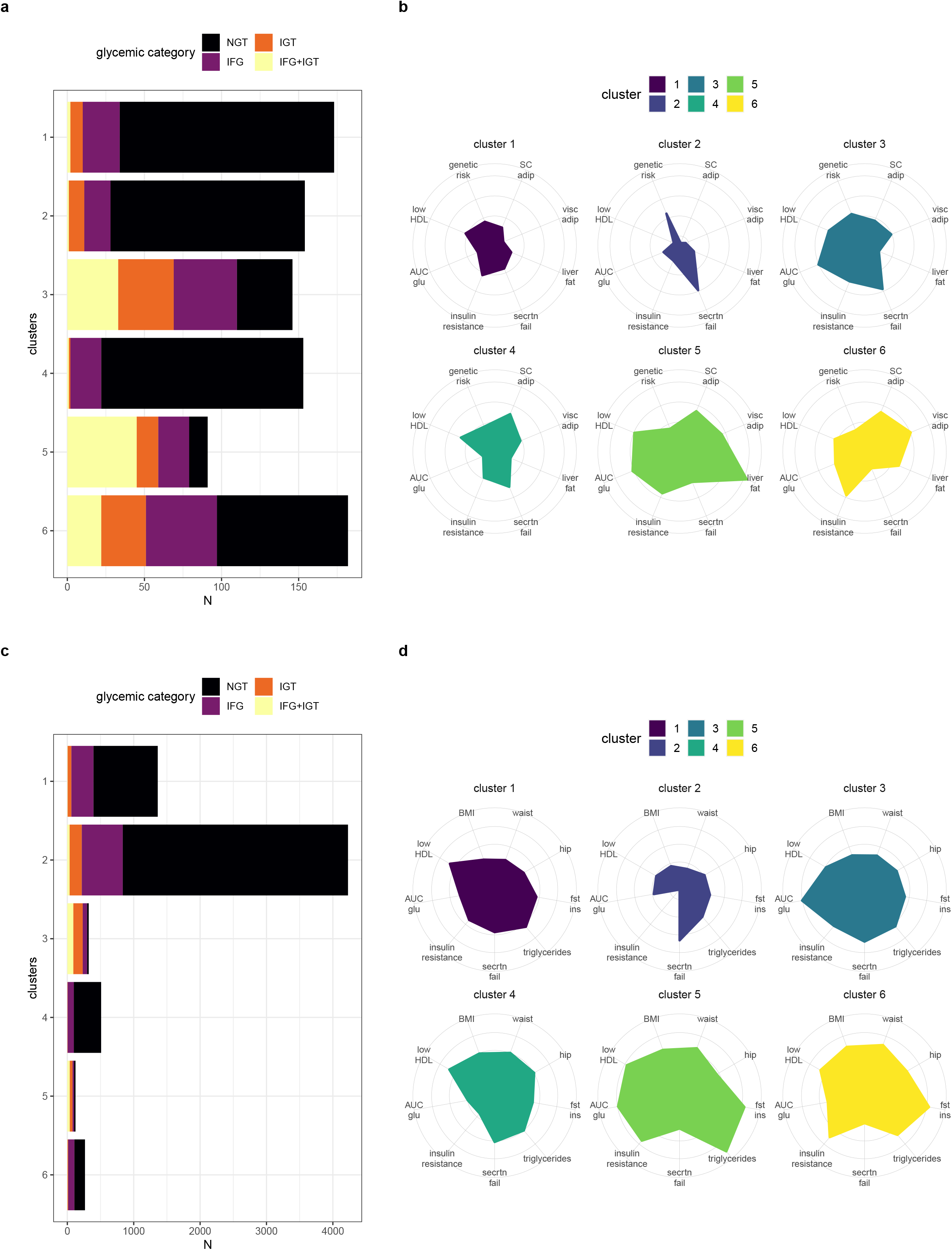
Distribution of the cluster feature variables. Partitioning of participants into 6 clusters along 8 variables in the TUEF/TULIP (N=899, Panel **a, b**) and 9 variables in the Whitehall-II cohort (N=6810, Panel **c, d**). Panel **a** and **c** show the number of participants in each cluster with colors indicating glycemic categories (NGT = normal glucose tolerance, IFG = impaired fasting glycaemia, IGT = impaired glucose tolerance, IFG+IGT concomitant impaired fasting glycaemia and impaired glucose tolerance). Panel **b** and **d** show the medoids (the representative subject, TUEF/TULIP) or the medians (Whitehall-II) of each cluster with the corresponding standardized level (Z-scores) of the feature variables. Clusters in the Whitehall-II cohort were identified using Euclidean distances from the median values of the proxy variables in TUEF/TULIP that have also been assessed in Whitehall-II. For the radar-charts (**b, d**), the Z-scores of insulin sensitivity, insulin secretion and HDL were directionally flipped (−1*Z-score) to yield polygon areas related to adverse variable effects.

There was a cluster-specific enrichment of the diabetes-related genetic variant rs10830963 in *MTNR1B* (ANOVA p=0.02 after Benjamini-Hochberg correction for multiple testing, Suppl.Table 4). Participants in cluster 3 had higher frequency of the diabetes-associated G allele compared with those in cluster 1 (uncorrected p=0.00036 for cluster 3 relative to cluster 1). Using the pathophysiological classification of diabetes-related genetic variants proposed by Udler et al^12^, we found differences within the beta-cell group (uncorrected p=0.001, p=0.007 after Benjamini-Hochberg correction, Figure 2.A). Pairwise comparisons showed significant differences between cluster 6 and each of clusters 1, 2 and 3 (ANOVA with Tukey’s post-hoc test p<0.05), suggesting a lower abundance of beta-cell function related risk alleles in cluster 6.

**Figure 2.**
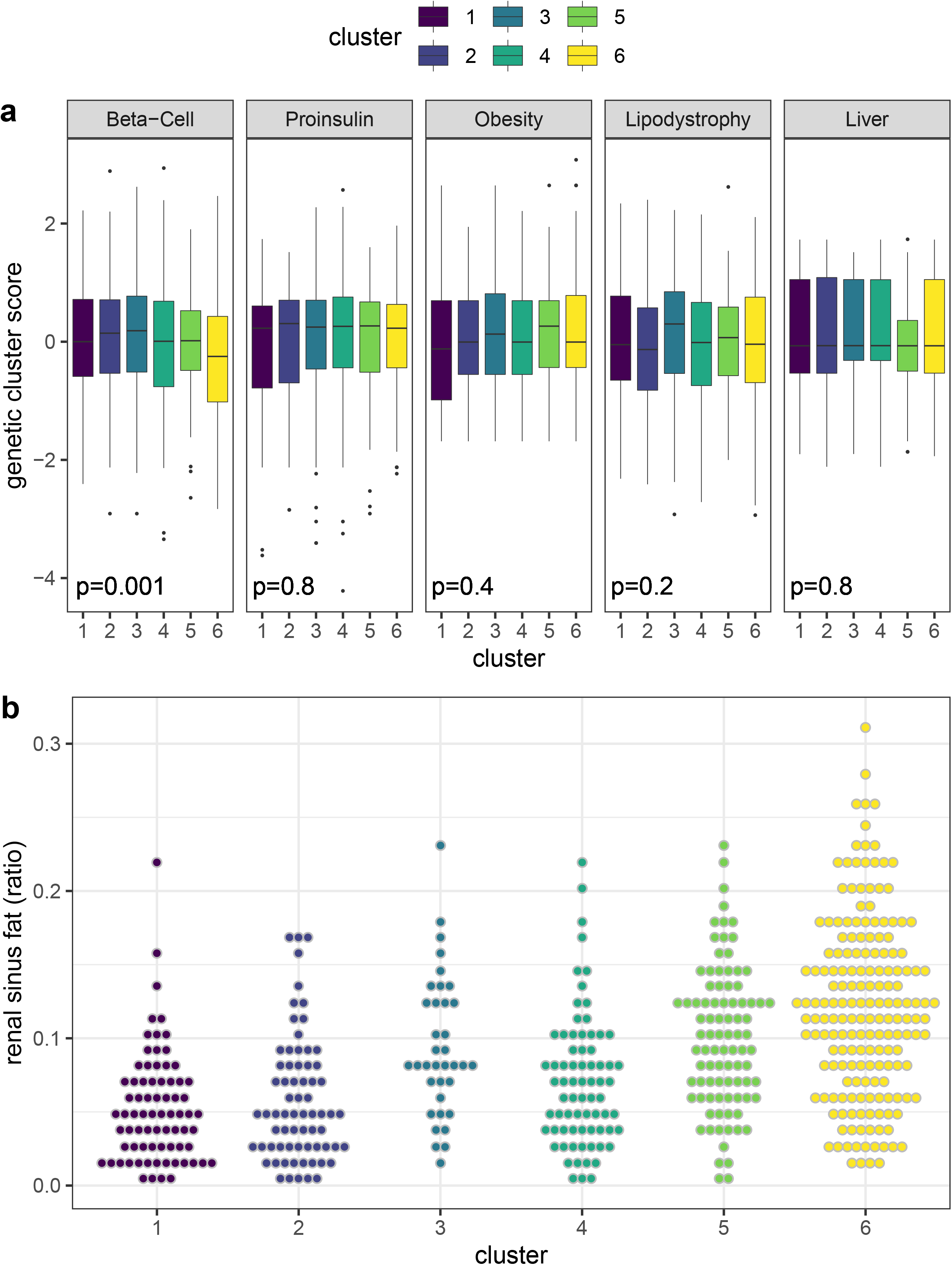
Characteristics potentially contributing to cluster pathomechanism. **a,** Mean pathway-specific genetic scores according to Udler et al across the 6 clusters of this work. Genetic scores (n=899 risk scores of individuals in TUEF/TULIP for each of the 5 specific pathways) were transformed to Z-scores to eliminate differences in absolute levels due to the differing number of genetic variants in each genetic pathway. Boxes (hinges) denote the 25^th^ and 75^th^ percentiles with an additional horizontal line indicating the median. Whiskers show the highest and lowest data points excluding outliers (defined as at least 1.5×interquartile range below the lower or above the upper hinge). Outliers are shown as individual data points. Differences were tested with one-way ANOVA. **b**, Distribution of renal sinus fat (ratio of sinus fat to kidney area, mean of left and right) for n=520 individuals with MRI-assessed renal sinus fat in TUEF/TULIP) across clusters (p=1.25×10^−26^ with one-way ANOVA). Pairwise tests for cluster 6 with Tukey’s test yielded the following p-values: p_5-6_ =0.02, p_3-6_=0.049, p_6-others_ <1×10^−14^.

In the longitudinal analysis, all participants with available data were followed for the development of diabetes, nephropathy, cardiovascular endpoints and all-cause mortality (Figure 3). The proportional hazards assessment in Whitehall II is shown in Suppl.Table 5. Diabetes incidence was the highest in cluster 5, followed by cluster 3 in both the TUEF/TULIP and Whitehall-II cohorts. Mean follow-up was 4.1 and 16.3 years, respectively. In TUEF/TULIP, participants in cluster 6 did not demonstrate an increased risk for diabetes (Figure 3.A). The diabetes-risk of cluster 6 was only moderately elevated in Whitehall II (HR 2.22[CI:1.7-2.89] compared with cluster 1. Cluster 3 and 5 showed hazard ratios of 3.45[CI:2.76-4.31] and 6.62[CI:5.06-8.67], respectively, compared with cluster 1, (Figure 3.C, Suppl.Table 5). By contrast, cluster 2 had a significantly lower risk of developing diabetes in the Whitehall II cohort compared with cluster 1 (HR 0.4[CI:0.33-0.47]). Current smoking was a risk factor for diabetes in Whitehall II, but did not affect the risk of diabetes for participants in clusters 3, 5 and 6 (Suppl.Table 6).

**Figure 3.**
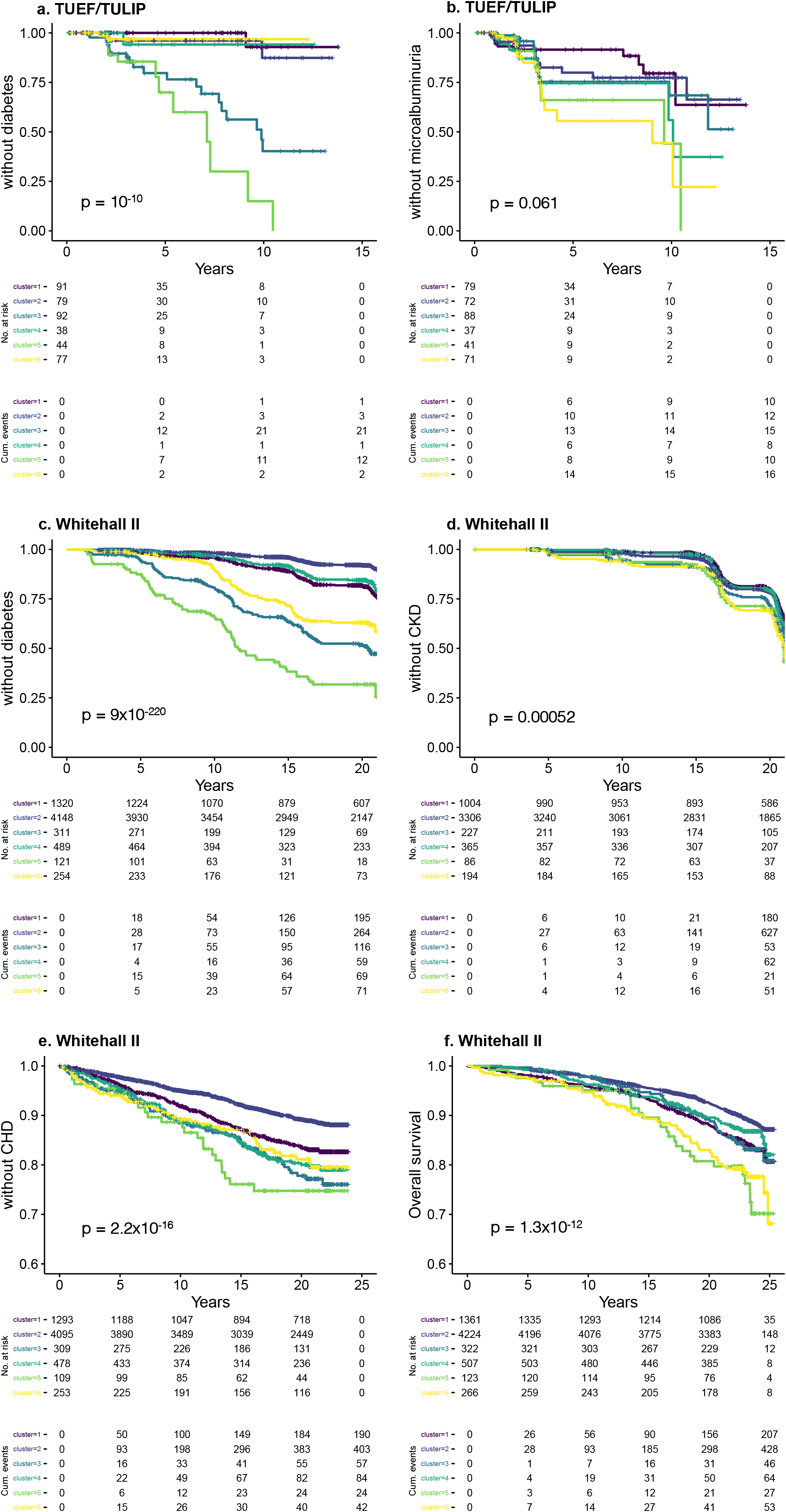
Cluster-specific outcomes. Kaplan-Meier curves showing cluster-specific probability of not developing diabetes (**a, c**), nephropathy (**c, d**) in the TUEF/TULIP and Whitehall-II cohorts, respectively. Cumulative probability of coronary heart disease (CHD, **e**) and overall mortality (**f**) are shown for the Whitehall II cohort. For diabetes incidence: n=421, mean follow-up 4.1 years, number of diabetes events = 40 in TUEF/TULIP and n=6643, mean follow-up 16.3 years, number of diabetes events = 828 in Whitehall II. For microalbuminuria incidence: n=388, mean follow-up 4.3 years, number of microalbuminuria events = 71 in TUEF/TULIP. In Whitehall II n=5182 mean follow-up 18.2 years with 1387 Stage 3 chronic kidney disease or worse (estimated glomerular filtration rate < 60 ml/min/1.73m^2^) incidences. For CHD, n=6537, mean follow-up 17.2 years, 800 events. For all-cause-mortality, n=6803, mean follow-up 21.1 years, 825 deaths. All p-values were computed with two-sided log-rank tests.

In Whitehall II, there were 201 participants with incident diabetes and a defined Ahqlvist diabetes classification^6^. Relatively few participants developed diabetes in the metabolically healthy clusters (cluster 1: 48 of 817 [5.9%], cluster 2: 62 of 2552 [2.4%], cluster 4: 14 of 314 [4.5%], out of those eligible for computation of the Ahlqvist-classes). Of these participants, most (34 of 48 [70.8%], 59 of 62 [95.2%] and 12 of 14 [85.7%], respectively) transitioned into mild diabetes classes according to the Ahlqvist-classification (mild obesity-related diabetes [MOD] and mild age-related diabetes [MARD]). 13 of 23 participants (57%) in cluster 6 (13 of 23 [57%]) developed severe insulin resistant diabetes (SIRD, Suppl.Table 7 and Extended Data 3).

We used two approaches to compare our multivariable clustering with glucose-based stratification alone. We first tested cumulative diabetes risk for the Hulman classes^13^ that are computed from the glucose course during an OGTT (Extended Data 4). Next, we stratified the baseline AUC glucose of Whitehall II into 5 quintiles, (Extended Data 5). In head-to-head comparisons, the cumulative diabetes risk of the high risk clusters 3 and 5 together was higher than that of Hulman-classes 3 and 4 together (p=0.04, TUEF/TULIP) and also higher than that of the top 2 AUC glucose quintiles (p<0.0001, Whitehall II, both log-rank tests). Thus, our cluster-based approach was superior to both of these approaches in delineating groups with high cumulative risk for development of diabetes.

The overall difference in the Kaplan-Meier curves for microalbuminuria did not reach statistical significance in TUEF/TULIP (mean follow-up 4.3 years, number of events=71, p_log-rank, uncorrected_=0.061, Figure 3.B). In the proportional hazard assessment, cluster 6 showed a significantly higher risk for microalbuminuria compared with cluster 1 (p=0.01). Results were similar but not significant for the Whitehall II participants with available baseline urine measurements (n=316, number of events=58, uncorrected p=0.058) when adjusting for baseline urinary albumin-to-creatinine ratio. In Whitehall II, participants in cluster 6 had a significantly higher risk for stage 3 chronic kidney disease or worse than cluster 1 (uncorrected p=0.0003, mean follow-up 18.2 years, number of events 1387, Figure 3.D, Extended Data 6). Individuals in the diabetes susceptible clusters 3 and 5 also demonstrated higher risks for chronic kidney disease relative to cluster 1 in Whitehall II (uncorrected p=0.004 and p=0.02, respectively, Suppl.Table 5). The fully adjusted model also controlled for smoking, cholesterol and triglycerides is shown in Suppl.Table 8. Given that participants in cluster 6 had elevated visceral fat, we hypothesized that this could be associated with fat in the renal sinuses, which is a risk factor for exercise-induced microalbuminuria^14^. TUEF/TULIP participants in cluster 6 had the most renal sinus fat compared with other clusters (p<0.05 for all pairwise comparisons, Tukey’s post-hoc test, Figure 2.B, N=199). It was higher than in cluster 5 after adjusting for potential confounders (Suppl.Table 9.A-D).

In the TUEF/TULIP cohort, we used carotid intima media thickness (IMT) as a proxy for cardiovascular end-points due to a lack of a register-based assessment of clinical events. IMT was associated with cluster membership (F=14.55, degrees of freedom=5, p<0.001). Each of clusters 3, 5 and 6 had higher IMT values than each of clusters 1, 2 or 4 (Extended Data 7 and Suppl.Table 1, p<0.002). After adjustment for sex, age, age^2^ and BMI, cluster 3 and 5 had higher IMT than cluster 1 (p<0.03). In the Whitehall II cohort, we evaluated the incidence of coronary heart disease (CHD, mean follow-up 17.2 years, 800 events, see Figure 2.E). As a combined vascular endpoint, we also investigated the incidence of CHD and stroke (mean follow-up 22.9 years, 1040 events, Suppl.Table 5). In the proportional hazard assessment, the elevated cardiovascular risk in cluster 5 was not independent from sex, age and BMI, but consistently lower in cluster 2 compared with cluster 1, also after adjustments (Suppl.Table 5). Compared with cluster 1 in Whitehall II, all-cause mortality was by about 40% higher for cluster 6 (Figure 3.F), while cluster 2 had a lower mortality rate, even after adjustments for covariates (Suppl.Table 5). The elevated mortality risk in cluster 6 (relative to cluster 1) was not affected by adjustment for smoking and lipids (full model in Suppl.Table 10).

## Discussion

The applied variable-based partitioning of individuals without type 2 diabetes yielded groups differing in risk for type 2 diabetes and its complications. We validated these findings using simple measures of the same pathophysiological constructs in a large occupational cohort.

Cluster 5 was identified as the subpopulation of the highest risk of type 2 diabetes, renal and vascular disease and all-cause mortality. Individuals in this cluster had obesity, insulin resistance, high levels of fatty liver and low insulin secretion. Cluster 6 represented an insulin resistant phenotype, in which participants had high amounts of visceral fat, but less liver fat and higher insulin secretion compared with cluster 5. About half of the participants in cluster 6 had prediabetes on enrollment in the TUEF/TULIP study. However, mean glycemia (AUC glucose) was lower than in cluster 5, and the risk of type 2 diabetes was considered to be moderate. Nonetheless, participants in cluster 6 had high risk for microalbuminuria and chronic kidney disease. Cardiovascular risk was not elevated in this cluster; however, overall mortality was about 40% higher than in the reference cluster 1 even after adjustment for confounders. Thus, clusters 5 and 6 both constitute obese, high-risk subpopulations with different glycemic, renal, cardiovascular and all-cause mortality risk profiles.Glucose does not seem to be the major driver of clinical events in cluster 6. Previous observations of an association of insulin resistance with diabetic nephropathy^15–17^ highlight insulin resistance as a probable underlying factor. The discrepancy between moderate type 2 diabetes and high nephropathy risk for cluster 6 is not dependent from baseline blood pressure. However, individuals in cluster 6 had elevated renal sinus fat, which could contribute to manifestation of nephropathy. We previously showed an association between renal sinus fat and exercise-induced albuminuria in a cross-sectional cohort and an association of microalbuminuria with renal sinus fat in individuals with non-alcoholic fatty liver disease^14,18^. In renal sinus fat and renal cell co-culture experiments, the combination of renal sinus fat and Fetuin-A induced inflammation indicate a combination of an insulin resistant metabolic milieu and adverse fat accumulation as a likely cause of organ damage^18^. This finding is consistent with the phenotypes of insulin resistance, moderately high liver fat and high renal sinus fat in cluster 6. Cluster 6, in which participants had moderate or delayed risk of diabetes, showed a relatively low genetic risk for type 2 diabetes and a low abundance of genetic variants from the beta-cell class in the Udler classification^12^. This result implies an effective compensation of insulin resistance through excellent beta-cell function. We speculate that hyperinsulinemia associated with the combination of good beta-cell function and insulin resistance contributes to renal disease and mortality^19–21^. Smoking was a risk factor both for diabetes and chronic kidney disease^22–24^, but did not explain the differences among clusters.

Contrast to the three high-risk clusters 3, 5 and 6, cluster 4 comprises participants with obesity but low glycemic deterioration. Phenotypic traits of individuals in this cluster are compatible with the concept of metabolically healthy obesity^25^. Cluster 4 was also associated with lower risk of type 2 diabetes, independently from sex, age, and BMI. Individuals in this cluster had body fat predominantly stored in subcutaneous rather than visceral depots, a pattern known to be metabolically more favorable^26^. In cluster 3, the partitioning identified a phenotype characterized by elevated genetic risk and low insulin secretion, which might explain the high diabetes incidence seen in this group. The moderately elevated visceral fat compartment correlates with pancreatic fat, which has been associated with disturbed insulin secretion in a prediabetic environment^18,27,28^. Cluster 3 with a disposition index as low as cluster 5, but higher insulin sensitivity could correspond to beta-cell dysfunction subphenotypes identified in previous studies^6,7,29^. Cluster 3 had high IMT, independent from sex, age and BMI. Increased cardiovascular risk was not replicated for this cluster in Whitehall II, but individuals in this cluster had a moderately elevated risk of chronic kidney disease.

Our clustering approach is not designed to provide definitive subphenotypes for individual patients in a clinical setting; however, the approach can be helpful for characterizing the metabolic heterogeneity prior to clinical manifestation of type 2 diabetes. The identification of such subphenotypes suggests some potential therapeutic implications. Individuals in cluster 5 are at imminent risk for diabetes and could benefit from high intensity dietary and/or lifestyle interventions aimed at weight loss and liver fat reduction. Individuals with the characteristics of cluster 3 might benefit from a standard aerobic exercise and dietary caloric restriction via reduction of visceral fat. Although clusters 3 and 5 have elevated genetic risk as non-modifiable risk factor, genetic predisposition might be protective against development of type 2 diabetes for individuals with a cluster 6 phenotype. This group could be easily overlooked when risk-stratification focuses on established diabetes-related glycemic cut-offs. Insulin resistance with or without prevalent prediabetes associates with renal disease and elevated mortality in cluster 6, which should motivate consideration of preventive measures even with low glycemic progression.

Our subphenotyping was performed in persons who did not yet suffer from diabetes, but who are at potentially increased risk, as demonstrated by the newly diagnosed cases in the follow-up period. The classification emerges partly from variables that require an OGTT. OGTT-derived glycemic traits can reasonably assess insulin sensitivity and secretion, particularly in the absence of diabetes. An elegant metabolic clustering of glycemic courses during OGTT has been proposed by Hulman et al^13^. We have applied an alternative approach with a broad set of variables in addition to OGTT. Our data complement other clustering approaches targeting the disentanglement of the heterogeneity of adult-onset diabetes^6,7,12^. We show that cluster 6 most strongly connects to the SIRD cluster of the Ahlqvist-classification^6,30^. Cluster 6 and SIRD bear similarities, such as an elevated risk of nephropathy in the absence of marked glucose elevation. Thus, accumulating data indicate that the pathogenesis of kidney damage in type 2 diabetes appears to be different from that of type 1 diabetes, with only a minor contribution of glycaemia in prediabetes and type 2 diabetes. Of note, by contrast with the Ahlqvist-classification, our work analyzed screen-detected diabetes cases as outcomes during the follow-up periods. These cases probably have milder phenotypes than clinically detected type 2 diabetes cases.

Our results are demonstrated in two independent study groups: a cohort by design enriched in diabetes-prone persons and a UK occupational cohort. This most likely contributes to the observed differences between the Kaplan-Meier plots in the two cohorts, especially for diabetes incidence. Given the lack of ethnic diversity of the investigated populations leveraged in our study, our findings might only be applicable to white European populations. We also acknowledge the limitations of the partitioning approach: there is uncertainty with regard to variable selection, the optimal number of clusters and whether these approaches are inferior to conventional predictions from multivariable modeling^29^. Additional specific limitations of our work are the different feature variable set and the moderate reassignment rate (63%) of the original clusters to the feature set of Whitehall II. Given the sophisticated nature of the variables in TUEF/TULIP cohort, the clinical utility of these features for metabolic classification could be limited. Further, in the TUEF/TULIP cohort, only about half of the population was available for follow-up visits. This high attrition rate could lead to a potential underestimation of the risk for diabetes and nephropathy in the TUEF/TULIP cohort. A final limitation is that the nephropathy models in Whitehall II are not adjusted for baseline eGFR due to a lack of baseline measurements and the absolute risks being low.

In summary, we show the feasibility of multi-variable subphenotyping in individuals without diabetes to disentangle metabolic heterogeneity prior to diagnosis of type 2 diabetes. The metabolic clusters identified here associate with future complications related to prediabetes, insulin resistance, future risk of type 2 diabetes and mortality. These subphenotypes likely reflect key pathologic features potentially underlying different fates of metabolic complications but are not aimed at classifying single patients in clinical practice; however, with further development and validation, such approaches could guide prevention and treatment strategies for cardiovascular and renal disease as well as type 2 diabetes.

## Data Availability

For TUEF/TULIP, all requests for data and materials will be promptly reviewed by the Data Access Steering Committee of the Institute of Diabetes and Metabolic Research, Tuebingen to verify if the request is subject to any intellectual property or confidentiality obligations. Individual level data may be subject to confidentiality. Any data and materials that can be shared will be released via a Material Transfer Agreement. Data access to individual-level data of the Whitehall II study is subject to a separate data sharing agreement according to the data sharing policy of Whitehall II. This policy conforms to the MRC Policy on Research Data Sharing. More details can be found on the Whitehall II webpage: https://www.ucl.ac.uk/epidemiology-health-care/research/epidemiology-and-public-health/research/whitehall-ii/data-sharing.

## Acknowledgments

We thank all the research volunteers for their participation. We thank all participants in the Whitehall II Study, Whitehall II researchers and support staff who made the study possible. We gratefully acknowledge the excellent technical assistance of the Diabetes Research Unit Diabetes Research and Metabolic Diseases of the Helmholtz Center Munich at the University of Tübingen, Germany.

We thank J. Kriebel and H. Grallert (Molecular Epidemiology, Helmholtz Center Munich) for generating the Global Screening Array data. This study was supported in parts by a grant (01GI0925) from the Federal Ministry of Education and Research (BMBF) to the German Center for Diabetes Research (DZD e.V.) and from the state of Baden-Württemberg to RW and AF (32-5400/58/2, Forum Gesundheitsstandort Baden-Württemberg). The UK Medical Research Council (MR/K013351/1; G0902037), British Heart Foundation (RG/13/2/30098), and the US National Institutes of Health (R01HL36310, R01AG013196) have supported collection of data in the Whitehall II Study.

## Author contributions

R.W. analyzed the data and wrote the manuscript. M.H., A.G.T., J.M., F.S., E.R., A.F. contributed to data acquisition, the interpretation of data and edited the manuscript. M.H.A., A.P. A.L.B and N.S contributed to the interpretation of data and edited the manuscript. H-U.H. and A.F. contributed to the concept of the work and edited the manuscript. All authors have reviewed the manuscript.

## Methods

### TUEF/TULIP cohort

Prediabetes subphenotyping was initially performed on a complete cases subset of participants of the Tuebingen Family study and Tuebingen Lifestyle Program (TUEF/TULIP)^2,3^, who had no missing values for the preselected phenotyping variables (N=899, baseline characteristics for this and the whole cohort are shown in the Suppl.Table 11). Participants were recruited from 2003 through 2018. Recruitment was mostly performed via newspaper announcements and e-mail bulletins. The studies have been designed to phenotype individuals at increased risk of diabetes. Eligibility criteria for inclusion comprised either a history of prediabetes, a family history of diabetes, a BMI greater than 27 kg/m^2^ or a history of gestational diabetes^2^. Participants underwent a frequently sampled OGTT and received MR-tomography-based measurement of body fat distribution and ^1^H-MR-spectroscopy-based measurements of hepatic fat content. Follow-up data was available for individuals who responded to invitations to follow-up appointments or participated in follow-up studies. The follow-up measurements were comparable to the initial assessments. Glycemic traits (fasting glucose, OGTT or HbA1c) were available for 421 participants, whereas urine sample during follow-up, for the determination of microalbuminuria, was available for 388 participants. The study protocol was approved by the Ethics Committee of the University of Tübingen (422/2002). All participants gave written informed consent.

### Whitehall II cohort

Data from the occupational Whitehall II cohort were accessed by a data sharing agreement. Details of the study have been described elsewhere^4^. In brief, the study was established to explore the relationship between socio-economic status, stress and cardiovascular disease. All London-based civil servants aged 33-55 years were invited in 1985-1988 and 10.308 (73%) participated. Since then, 5 further clinical examinations have taken place that are available for data sharing at approximately 5-year intervals (phases 3,5, 7, 9 and 11). The study was approved by the Joint UCL/UCLH Committees on the Ethics of Human Research (Committee Alpha). For the current analysis, the baseline was defined as the first available fasting OGTT (>=8 hours of fasting for morning and >=5 hours of fasting after a light fat-free breakfast eaten before 8 am for afternoon OGTTs). Participants with prevalent or incident diabetes at baseline and those with non-white ethnicities were excluded. From the 6916 available baseline OGTTs, 6810 were complete cases in regards of the used clustering variables und underwent cluster assignments. The cohort characteristics are reported in Suppl.Table 12.

### Variable selection and de novo clustering in TUEF/TULIP

We aimed to identify subphenotypes that reflect differences in pathophysiological processes in the natural history of type 2 diabetes. The main paradigm of type 2 diabetes pathogenesis is an insufficient compensatory increase of insulin secretion in response to insulin resistance^31^. Therefore, insulin sensitivity and insulin secretion are key variables^6,7^. We used OGTT-based indices of insulin sensitivity (Matsuda-index)^32^ and insulin secretion (AUC_0-30_ C-peptide/AUC_0-30_ glucose) that correlate well with gold-standard measures and are preferable to static measurements obtained in the fasting state^33,34^. Glycaemia was quantified in the partitioning procedure as AUC_0-120_ glucose. Furthermore, we aimed to capture diverse etiologies of insulin resistance by accounting for visceral and subcutaneous adipose tissue volume (VAT and SCAT), that have distinct metabolic characteristics^35^. We especially focused on elevated liver fat content, as it is strongly associated with insulin resistance^36^. HDL-cholesterol levels have been long known as explanatory variables of the metabolic syndrome and insulin resistance^37^. Moreover, causal inference from large genomic datasets provides evidence not only for a genetic correlation of HDL-cholesterol levels with type 2 diabetes, but also for a causal link between HDL-cholesterol levels and type 2 diabetes^9^. We also added a genome-wide polygenic risk score (PRS) to the analysis to better differentiate between genetically determined beta-cell dysfunction and environmentally determined beta-cell dysfunction. The correlation of the clustering variables is reported in Suppl.Table 13.

For computation of the PRS, we used the LDpred algorithm of Vilhjalmson *et al*.^38^ on a combination of BMI-adjusted effect sizes and p-values from a meta-analysis in ∼900.000 European individuals and genotypes^11^. After quality control, exclusion of multi-allelic and low-frequency variants, we combined 484.788 variants from the two datasets, yielding an estimated genome-wide SNP-heritability of 0.069. Of the top 94 diabetes-related genetic variants shown in the latest large-scale genome-wide association study^11^, 63 were genotyped in TUEF/TULIP. The association of cluster-assignment with the genotype was tested separately for each variant using ANOVA to analyze the enrichment of certain genotypes in clusters. A further genetic-pathophysiologic classification of clusters was performed according to data from Udler et al^12^. Here, we computed the genetic risk score for every individual and every genetic class (beta-cell, proinsulin, obesity, lipodystrophy and liver/lipid) taking only weights >= 0.75 into account, as described in the original publication. The classification of glucose response curves according to Hulman et al (Hulman-classes) was performed with the corresponding web-calculator from 5-point OGTT glucose values in the TUEF/TULIP study^13^.

### Cluster assignment in the Whitehall II cohort

For assigning participants in the Whitehall II cohort to clusters established in TUEF/TULIP, we used proxy variables. Since liver fat, visceral adipose tissue and subcutaneous adipose tissue were not available in the Whitehall II cohort, and only two-point OGTTs were performed, other anthropometric variables and analytes were employed instead of these variables. Variables were selected based on statistical consideration (correlation) and pathophysiologic (theoretical) connection to the original trait (e.g. liver fat – fasting triglycerides, fasting insulin and waist circumference). Transaminase activity was not available during the early phases of the Whitehall II study. The final variable set was selected upon the highest agreement in re-identification of the original cluster assignments using the new proxy variables in TUEF. The variables used in Whitehall II comprised glycemia during glucose challenge, insulin sensitivity^32^, Stumvoll’s first phase insulin secretion index using insulin and glucose levels at fasting and at 120 min during OGTT^39^, fasting insulin, fasting triglycerides, waist circumference, hip circumference, BMI and HDL-cholesterol. The median values of these variables in TUEF/TULIP were used to assign participants to clusters in Whitehall II (Extended data 1) by taking the nearest neighbors of the 6 cluster-centers based upon Euclidean distances. Since Whitehall-II used a restricted CVD-focused genotyping platform with only 48000 markers and the release of full-scale genotyping data was not readily available, we decided to omit the genetic risk score from the re-assignment procedure. Despite these limitations, successful re-assignment of the clusters was achieved in 63% of the original TUEF cohort.

### OGTT and laboratory analysis

All participants of TUEF/TULIP received a 75-g glucose solution (Accu-Check Dextro, Roche) at 8 a.m. following an overnight fast. Venous blood was obtained through an indwelling venous catheter before and 30, 60, 90 and 120 minutes after glucose ingestion. In the Whitehall II cohort, the OGTT procedure has been described earlier. In short, venous blood samples were collected after an overnight fast in the morning (≥8 hours of fasting) or in the afternoon after no more than a light fat-free breakfast eaten before 08.00 h (≥5 hours of fasting) followed by a standard 75g OGTT with a venous blood sample taken 2 hours after ingestion of the glucose solution. Glucose was analyzed in the Whitehall II study using an YSI glucose analyser (Yellow Springs Instruments). Glucose values were measured in TUEF/TULIP directly using a bedside glucose analyzer (YSI, Yellow Springs, CO or Biosen C-line, EKF-diagnostic, Barleben). In TUEF/TULIP, all other obtained blood samples were put on ice, the serum was centrifuged within 2 hours. Plasma insulin and C-peptide were determined by an immunoassay with the ADVIA Centaur XP Immunoassay System and HDL was measured using the ADVIA XPT clinical chemical analyser (all from Siemens Healthineers, Eschborn, Germany), while triglycerides were measured with standard colorimetric methods using a Bayer analyzer. In Whitehall II, insulin was measured with an in-house human insulin RIA and later with a DAKO ELISA kit (DAKO Cytomatin Ltd, Ely, UK). Serum creatinine was measured using a kinetic colorimetric (Jaffe) method on a Roche “P” Modular system (phase 9) and on a COBAS 8000 system (phase 11). Lipid measurements were described previously^40^. HbA1c measurements were performed using Tosoh glycohemoglobin analyzers in both studies (Tosoh Bioscience Tokyo Japan).

### Body fat distribution, liver fat content and renal sinus fat

Body fat distribution variables, i.e., VAT and SCAT, were determined by whole-body T1-weighted MRI as described earlier^41^. Liver fat content was measured by volume selective ^1^H-MR spectroscopy^42^. Renal sinus fat was measured with manual segmentation from MR image slices specifically in cluster 5 and 6 using a method described previously^14^. The operator performing the segmentation (JM) was not aware of the cluster assignments. The procedure could not be completed in 6 participants (2% missing) due to breathing artefacts in the images. Renal sinus fat data for clusters 1 to 4 were partly available from segmentations for previous projects (mean data availability 40% over cluster 1 to 4).

### Outcomes

For detection of incident diabetes, either of the following was used: clinically ascertained diabetes (from patient history, or by the use of a diabetes-medication), an elevated fasting glucose (>=7 mmol/l), post-challenge glucose (>=11.1. mmol/l, or HbA1c (48 mmol/mol or 6.5%) in both cohorts. To assess the Ahlqvist-classification^6^ for the subtypes of diabetes in Whitehall II, we used insulin-based HOMA2-indices, because C-peptide was not measured. GAD measurements were not available. HbA1c assessment had been introduced beginning with Phase 7. Cluster assignment was performed using the lowest Euclidean distances from the published cluster centers in the All New Diabetes in Scania (ANDIS) cohort after scaling the variables for the means and SDs of the ANDIS cohort. Microalbuminuria was assessed in TUEF/TULIP upon the first occurrence from morning spot urine using the albumin-to-creatinine ratio (ACR). Measurements with excessive leukocyturia (175 measurements out of 3218) were excluded from this analysis. Microalbuminuria was established with an ACR>=30 mg/g creatinine. Carotid intima-media thickness (IMT), which is associated with future cardiovascular and cerebrovascular events^43^, was determined by a high-resolution ultrasound of the left and right common carotid artery. A trained physician who was unaware of the clinical and laboratory variables of the participants performed B-mode ultrasound imaging using a linear ultrasound transducer (10-13 MHz; AU5 Harmonic, ESAOTE BIOMEDICA, Hallbergmoos, Germany). IMT was specified according to the European Mannheim carotid intima-media thickness consensus criteria^44^. To ascertain renal disease, we used estimated glomerular filtration rate calculated using the CKD-EPI creatinine equation^45^. Serum creatinine was available from phase 9. Only participants with at least one eGFR value went into these analyses. Stages of chronic kidney disease were ascertained with the Kidney Disease: Improving Global Outcomes (KDIGO) classification^46^. Ascertainment of coronary heart disease and mortality in Whitehall-II has been described earlier^47^. In brief, incident CHD was defined as CHD death, nonfatal CHD and typical angina ascertained from clinical records, without self-reported cases from the Rose angina questionnaire. The cases were ascertained from participants’ general practitioners, information extracted from hospital medical records by study nurses, or data from the NHS Hospital Episode Statistics (HES) and death register databases obtained after linking the participants’ unique NHS identification numbers to this national database. Mortality data until June 2015 was drawn from the British National Mortality Register (National Health Service [NHS] Central Register) using each participants’ NHS identification number.

### Statistical analysis

Statistical analyses were performed using R version 3.4.3^48^. In the clustering analysis, distances were computed as Gower-distances using standardized variables (scaled to a mean of 0 and SD of 1). Participants with outlier variables (absolute standardized levels >= 5) were excluded from the clustering procedure. To find the optimal cluster count, we evaluated the dendogram and silhouette-widths. The clustering procedure was performed with the partitioning around medoids (pam) method in the R-package “cluster”, which is a more robust version of k-means clustering^49^. Using repeated subsetting with the clusterboot function from the fpc package, the mean Jaccard-similarity measure was 0.74 across all clusters.^50^ To further validate the stability of clusters, we iterated the clustering procedure for each of the 429 participants who had repeated measurements comprising all clustering variables (mean number of measurements 2.6±0.9, follow-up duration 4.2±3.6 years, also see Extended Data 8). We assessed the per-participant agreement of the generated 1112 cluster assignments using interrater reliabilities. The ICC2k value for cluster agreement was 0.72 (CI 0.68 – 0.76). Detailed reports on means and SDs of the clustering variables in both cohorts and the cluster medians are provided in Suppl.Tables 14-15.

Cluster means were compared using ANOVA. Specific outcomes were compared using ANCOVA adjusting for covariates such as sex, age and BMI. Post-hoc comparisons were performed using Tukey’s honest significant differences procedure. Endpoints related to diabetes complications were analyzed in the follow-up data of both cohorts using survival analysis and proportional hazard models. Differences in cumulative risks for reaching endpoints were tested with log-rank tests. When not indicated otherwise, the uncorrected p-value of a specific cluster’s risk relative to cluster 1 is provided in the proportional hazard analysis. Given the relatively low number of outcomes in TUEF/TULIP (40 for diabetes and 71 for microalbuminuria), assessment of proportional hazards adjusted for potential confounders was performed in the Whitehall II cohort only. Proportional hazards assumptions were tested by visualization of the Schoenfeld-residuals. The performed statistical tests were two-sided.

## Data availability

For TUEF/TULIP, all requests for data and materials will be promptly reviewed by the Data Access Steering Committee of the Institute of Diabetes and Metabolic Research, Tübingen to verify if the request is subject to any intellectual property or confidentiality obligations. Individual level data may be subject to confidentiality. Any data and materials that can be shared will be released via a Material Transfer Agreement. Data access to individual-level data of the Whitehall II study is subject to a separate data sharing agreement according to the data sharing policy of Whitehall II. This policy conforms to the MRC Policy on Research Data Sharing. More details can be found on the Whitehall II webpage: https://www.ucl.ac.uk/epidemiology-health-care/research/epidemiology-and-public-health/research/whitehall-ii/data-sharing.

## Code availability

The R code used to generate all results of this manuscript is available upon request. Requests will be reviewed by the Data Access Steering Committee of the Institute of Diabetes and Metabolic Research, Tübingen.

