## Extended Data and Supplementary Tables for "Pathophysiology-based subphenotyping of individuals at elevated risk for type 2 diabetes"

#### Extended Data 1

Assignment of proxy variables from the Whitehall II cohort (variables that were both assessed in Whitehall-II and the original clustering cohort TUEF/TULIP) to the original clustering variables. Clusters were identified in Whitehall II using the Euclidean distances of the subjects computed from these variables to the cluster medians in TUEF/TULIP. The upper row shows the original clustering variables available in TUEF/TULIP, the lower row the variables in Whitehall-II. Arrows show the physiological connection between the variables.

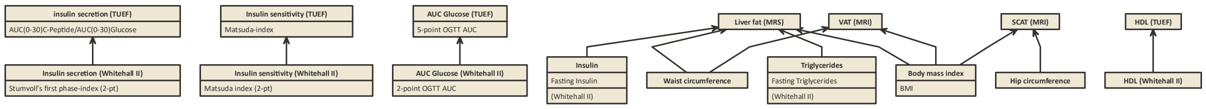

#### Extended Data 2

Key features of the clusters

| Cluster | Main feature | Obesity and fat distribution | Insulin sensitivity | Insulin secretion | Glycemia | Other specific features |
|---|---|---|---|---|---|---|
| 1 | Low risk | Overweight | Average | Adequate | Mostly NGT | |
| 2 | Very low risk | Normal | Good | Adequate | Mostly NGT | |
| 3 | Beta cell failure | Overweight/ Obese | Moderately low | Low | Mostly prediabetes | Increased genetic T2D risk |
| 4 | Low risk obese | Obese | Good | Adequate | Mostly NGT | |
| 5 | High risk insulin resistant fatty liver | Obese | Very low | Low | Mostly prediabetes (most of the latter IGT with or without IFG) | Above average genetic T2D risk, very high liver fat |
| 6 | High risk visceral fat nephropathy | Obese | Low | Moderately low | NGT and prediabetes (most of the latter IFG) | Low genetic T2D risk, high visceral fat, high renal sinus fat |

NGT: normal glucose tolerance, IFG: impaired fasting glucose, IGT: impaired glucose tolerance, T2D: type 2 diabetes

#### Extended Data 3

Transitions into Ahlqvist-diabetes-classes (right hand side) for subjects who were assigned to clusters in the Whitehall II study and developed diabetes during follow-up (left hand side, N=201).

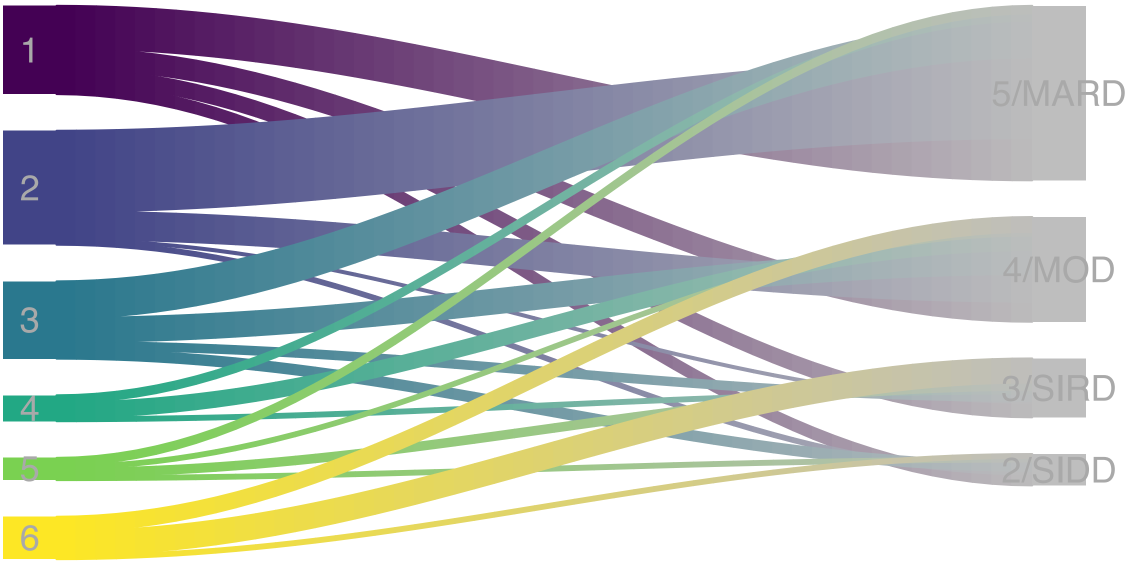

#### Extended Data 4.

Kaplan-Meier curves to compare the risk discrimination between Hulman-classes (A, n=416 individuals with follow-up) and clusters (B, n=421 individuals with follow-up) showing probabilities of remaining diabetes free in the TUEF/TULIP cohort. P-values indicate two-sided log-rank tests.

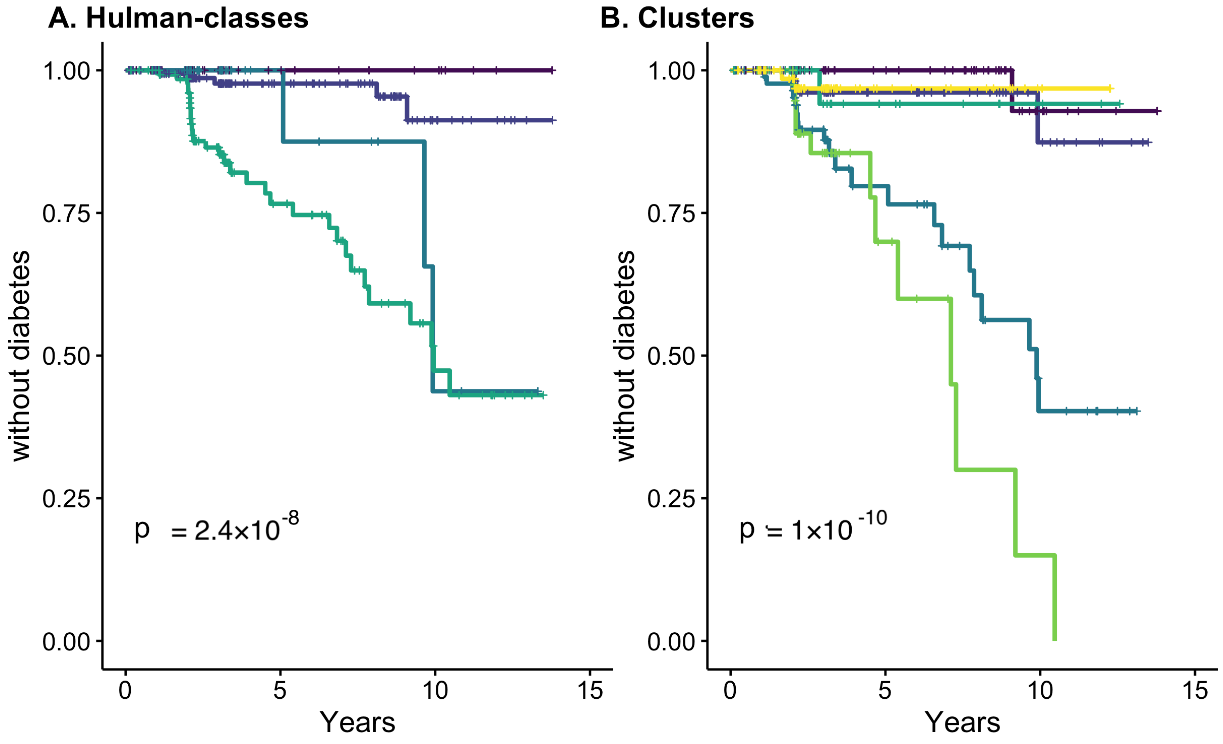


**A. No. at risk**

| | 0 | 5 | 10 | 15 |
|---|---|---|---|---|
| hulman_class=class1 | 55 | 11 | 6 | 0 |
| hulman_class=class2 | 186 | 59 | 13 | 0 |
| hulman_class=class3 | 38 | 8 | 2 | 0 |
| hulman_class=class4 | 137 | 41 | 11 | 0 |

**B. No. at risk**

| | 0 | 5 | 10 | 15 |
|---|---|---|---|---|
| cluster=1 | 91 | 35 | 8 | 0 |
| cluster=2 | 79 | 30 | 10 | 0 |
| cluster=3 | 92 | 25 | 7 | 0 |
| cluster=4 | 38 | 9 | 3 | 0 |
| cluster=5 | 44 | 8 | 1 | 0 |
| cluster=6 | 77 | 13 | 3 | 0 |

**A. Cum. events**

| | 0 | 5 | 10 | 15 |
|---|---|---|---|---|
| hulman_class=class1 | 0 | 0 | 0 | 0 |
| hulman_class=class2 | 0 | 3 | 5 | 5 |
| hulman_class=class3 | 0 | 0 | 3 | 3 |
| hulman_class=class4 | 0 | 21 | 31 | 32 |

**B. Cum. events**

| | 0 | 5 | 10 | 15 |
|---|---|---|---|---|
| cluster=1 | 0 | 0 | 1 | 1 |
| cluster=2 | 0 | 2 | 3 | 3 |
| cluster=3 | 0 | 12 | 21 | 21 |
| cluster=4 | 0 | 1 | 1 | 1 |
| cluster=5 | 0 | 7 | 11 | 12 |
| cluster=6 | 0 | 2 | 2 | 2 |

#### Extended Data 5.

Kaplan-Meier curves to compare the risk discrimination between quintiles of baseline glucose AUC levels (A, n=6643 individuals with follow-up) and clusters (B, n=6643 individuals with follow-up) for diabetes development in the Whitehall II study. P-values indicate two-sided log-rank tests.

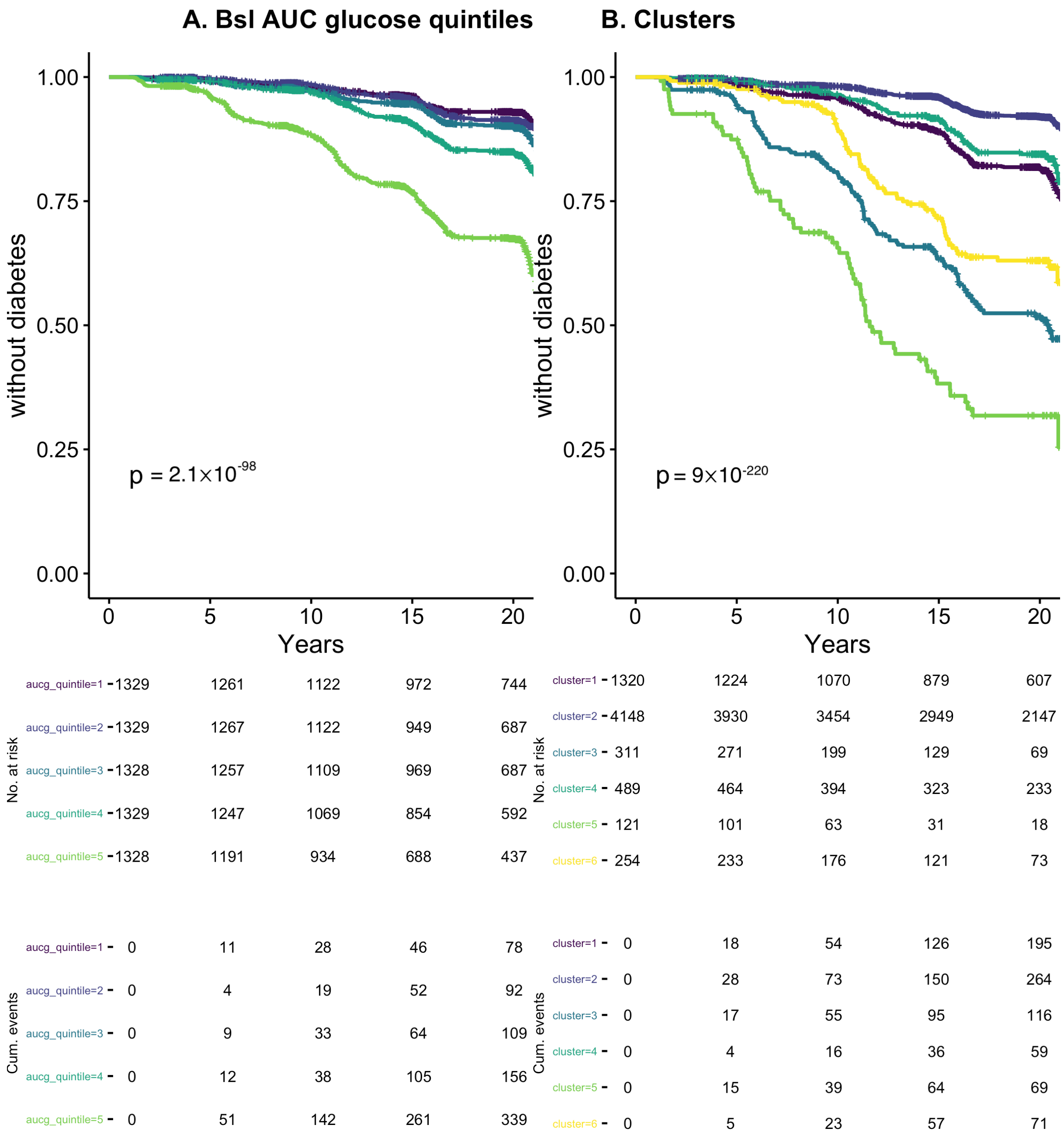

#### Extended Data 6

Cumulative incidence of chronic kidney disease stage 3 or worse in the Whitehall-II study (N=5182)

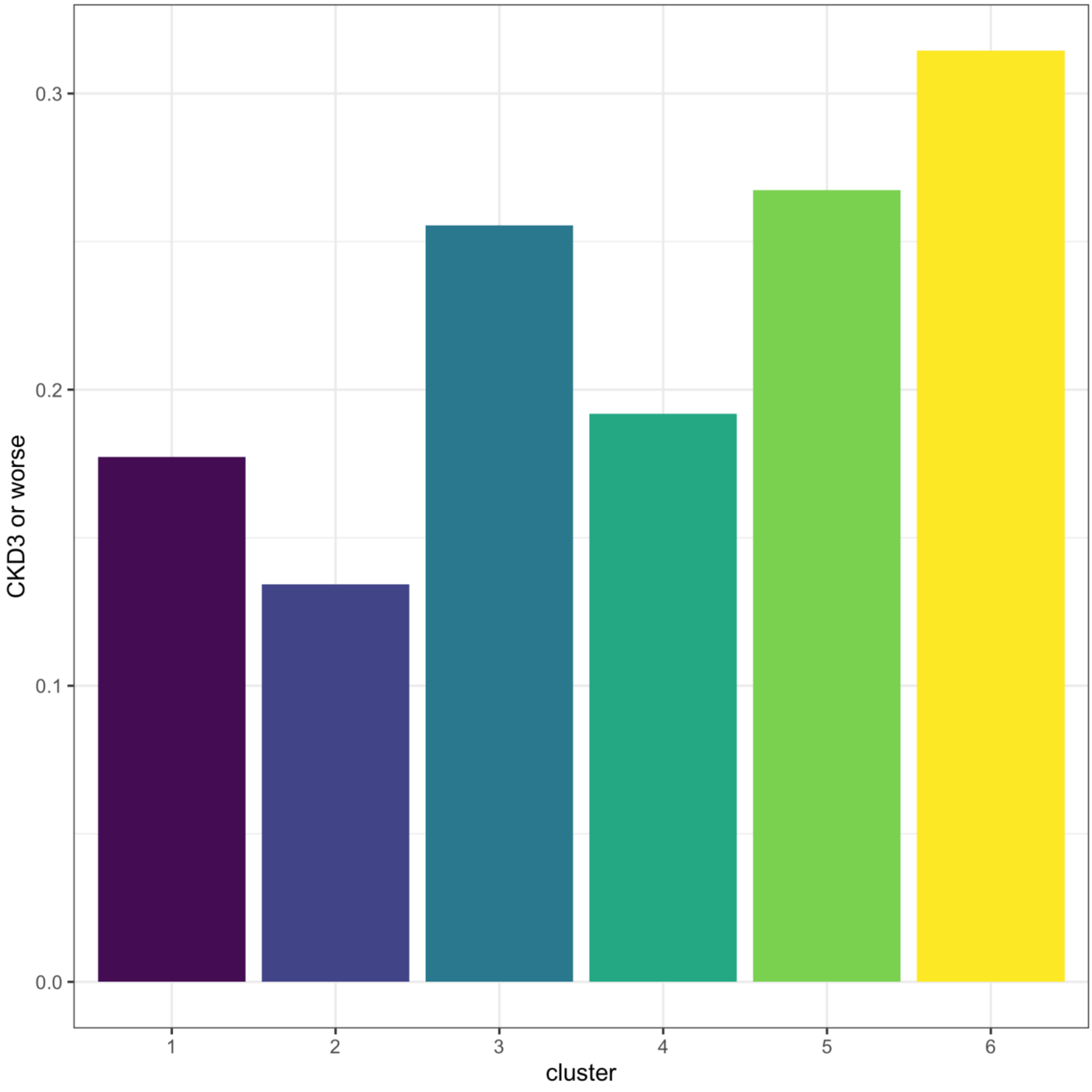

Extended Data 7.

Cluster-stratified carotid intima media thickness (IMT) in the subset with ultrasound measurements (N=479) in the TUEF/TULIP study. IMT was measured in 60%, 37%, 46%, 72%, 45% and 55% of the participants of cluster 1 through 6, respectively. Boxes (hinges) denote the 25th and 75th percentiles with an additional horizontal line indicating the median. Whiskers show the highest and lowest data points excluding outliers (defined as at least 1.5×interquartile range below the lower or above the upper hinge). Outliers are shown as individual data points.

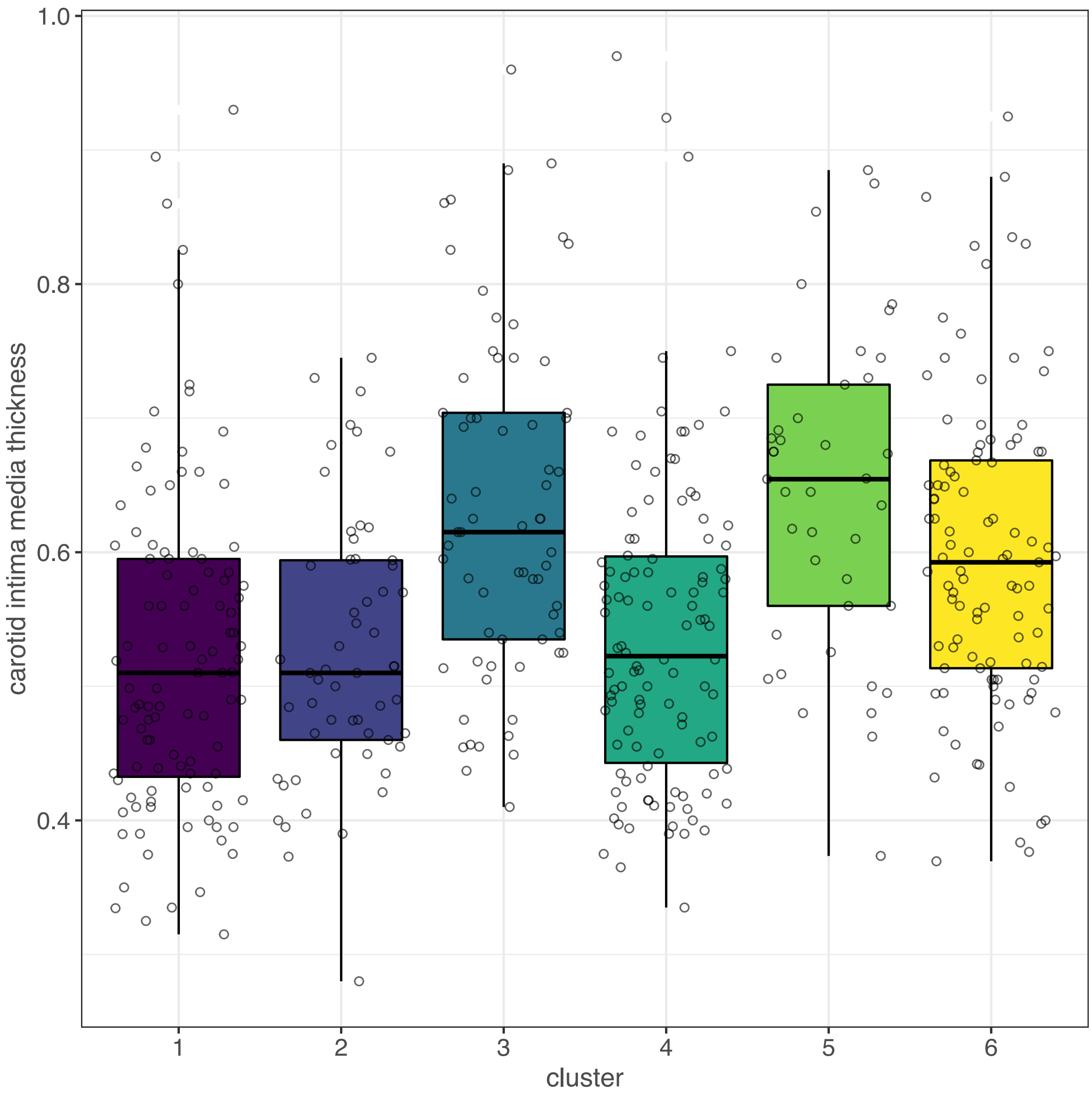

#### Extended Data 8

Cluster stability plot showing all consecutive cluster transitions in the iterative re-clustering of repeated measurements in TUEF/TULIP (N=429 individuals with repeated measurements)

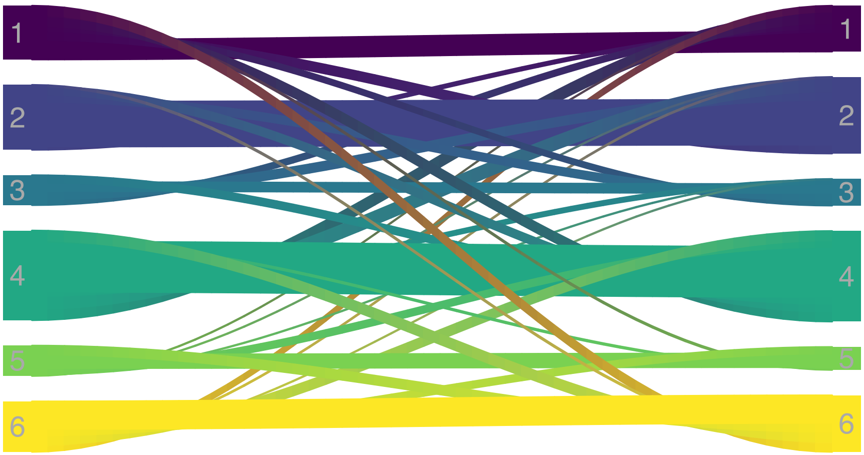

### Supplementary Information

Pathophysiology-based subphenotyping of individuals at elevated risk for type 2 diabetes

### Supplementary Tables

#### Supplementary Table 1.

Post-hoc tests for comparison of cluster means in the **TUEF/TULIP** study. Numbers denote p-values for all pairwise comparisons computed with Tukey's test (n=899 individuals).

| variable | **2-1** | **3-1** | **3-2** | **4-1** | **4-2** | **4-3** | **5-1** | **5-2** | **5-3** | **5-4** | **6-1** | **6-2** | **6-3** | **6-4** | **6-5** |
|---|---|---|---|---|---|---|---|---|---|---|---|---|---|---|---|
| age | 0.37 | $<1\times10^{-14}$ | $8.63\times10^{-12}$ | 0.97 | 0.87 | $<1\times10^{-14}$ | $8.57\times10^{-10}$ | $2.11\times10^{-05}$ | 0.65 | $1.19\times10^{-07}$ | $6.82\times10^{-09}$ | $5.64\times10^{-04}$ | 0.0058 | $2.14\times10^{-06}$ | 0.68 |
| BMI (kg/m2) | $1.27\times10^{-12}$ | $5.12\times10^{-06}$ | $<1\times10^{-14}$ | $<1\times10^{-14}$ | $<1\times10^{-14}$ | $5.13\times10^{-06}$ | $<1\times10^{-14}$ | $<1\times10^{-14}$ | $<1\times10^{-14}$ | $9.36\times10^{-07}$ | $<1\times10^{-14}$ | $<1\times10^{-14}$ | $<1\times10^{-14}$ | $3.80\times10^{-13}$ | 0.93 |
| waist circumference (cm) | $3.15\times10^{-09}$ | $8.18\times10^{-11}$ | $3.23\times10^{-13}$ | $3.74\times10^{-13}$ | $3.23\times10^{-13}$ | 0.6 | $3.23\times10^{-13}$ | $3.23\times10^{-13}$ | $2.45\times10^{-12}$ | $1.39\times10^{-08}$ | $3.23\times10^{-13}$ | $3.23\times10^{-13}$ | $3.75\times10^{-13}$ | $1.78\times10^{-11}$ | 1 |
| hip circumference (cm) | $1.44\times10^{-06}$ | 0.003 | $3.75\times10^{-13}$ | $3.79\times10^{-13}$ | $3.23\times10^{-13}$ | $7.89\times10^{-08}$ | $3.78\times10^{-13}$ | $3.23\times10^{-13}$ | $6.37\times10^{-11}$ | 0.38 | $3.23\times10^{-13}$ | $3.23\times10^{-13}$ | $3.80\times10^{-13}$ | $7.48\times10^{-04}$ | 0.68 |
| total adipose tissue MRI (liter) | $1.67\times10^{-09}$ | $5.00\times10^{-06}$ | $<1\times10^{-14}$ | $<1\times10^{-14}$ | $<1\times10^{-14}$ | $<1\times10^{-14}$ | $<1\times10^{-14}$ | $<1\times10^{-14}$ | $<1\times10^{-14}$ | 0.031 | $<1\times10^{-14}$ | $<1\times10^{-14}$ | $<1\times10^{-14}$ | $3.73\times10^{-07}$ | 0.54 |
| sq adipose tissue MRI (liter) | $4.35\times10^{-06}$ | 0.0011 | $8.92\times10^{-14}$ | $4.22\times10^{-14}$ | $3.83\times10^{-14}$ | $2.38\times10^{-11}$ | $3.83\times10^{-14}$ | $3.83\times10^{-14}$ | $9.04\times10^{-14}$ | 0.088 | $3.83\times10^{-14}$ | $3.83\times10^{-14}$ | $3.83\times10^{-14}$ | $1.39\times10^{-07}$ | 0.21 |
| visceral adipose tissue MRI (liter) | 0.036 | $<1\times10^{-14}$ | $<1\times10^{-14}$ | $6.16\times10^{-09}$ | $<1\times10^{-14}$ | 0.43 | $<1\times10^{-14}$ | $<1\times10^{-14}$ | $2.15\times10^{-08}$ | $1.40\times10^{-13}$ | $<1\times10^{-14}$ | $<1\times10^{-14}$ | $1.47\times10^{-10}$ | $<1\times10^{-14}$ | 1 |
| sq to visceral adipose ratio | 0.49 | $9.11\times10^{-08}$ | 0.0011 | 0.98 | 0.15 | $4.16\times10^{-09}$ | $2.41\times10^{-06}$ | 0.0039 | 1 | $1.66\times10^{-07}$ | $2.30\times10^{-05}$ | 0.051 | 0.77 | $1.25\times10^{-06}$ | 0.79 |
| visceral adipose % of total | 1 | $3.03\times10^{-09}$ | $9.51\times10^{-10}$ | 1 | 0.96 | $1.47\times10^{-07}$ | $8.74\times10^{-06}$ | $3.00\times10^{-06}$ | 0.99 | $1.03\times10^{-04}$ | $1.18\times10^{-05}$ | $3.73\times10^{-06}$ | 0.49 | $2.43\times10^{-04}$ | 0.93 |
| liver fat content | 0.11 | 0.0024 | $2.05\times10^{-08}$ | 0.99 | 0.029 | 0.024 | $<1\times10^{-14}$ | $<1\times10^{-14}$ | $<1\times10^{-14}$ | $<1\times10^{-14}$ | $<1\times10^{-14}$ | $<1\times10^{-14}$ | $<1\times10^{-14}$ | $<1\times10^{-14}$ | $<1\times10^{-14}$ |
| renal sinus fat (mean of r&l, %) | 0.98 | $6.53\times10^{-04}$ | 0.0067 | 0.18 | 0.58 | 0.24 | $3.70\times10^{-08}$ | $3.79\times10^{-06}$ | 0.99 | 0.005 | $2.07\times10^{-10}$ | $2.07\times10^{-10}$ | 0.05 | $3.42\times10^{-10}$ | 0.023 |
| diastolic blood pressure (mmHg) | 0.76 | 0.0042 | $2.74\times10^{-05}$ | 0.98 | 0.34 | 0.054 | $2.14\times10^{-13}$ | $2.02\times10^{-13}$ | $1.17\times10^{-05}$ | $2.82\times10^{-12}$ | $7.29\times10^{-08}$ | $4.93\times10^{-11}$ | 0.35 | $1.09\times10^{-05}$ | 0.0075 |
| systolic blood pressure (mmHg) | 0.62 | $6.68\times10^{-06}$ | $3.10\times10^{-09}$ | 1 | 0.63 | $1.67\times10^{-05}$ | $2.36\times10^{-13}$ | $2.00\times10^{-13}$ | 0.0049 | $3.33\times10^{-13}$ | $1.12\times10^{-09}$ | $2.91\times10^{-13}$ | 0.86 | $5.52\times10^{-09}$ | 0.072 |
| heart rate (BPM) | 0.64 | 1 | 0.71 | 0.91 | 1 | 0.94 | $1.79\times10^{-04}$ | $5.76\times10^{-07}$ | $2.95\times10^{-04}$ | $4.89\times10^{-06}$ | 0.11 | $9.97\times10^{-04}$ | 0.13 | 0.0064 | 0.19 |
| fasting glucose (mmol/l) | 0.7 | $<1\times10^{-14}$ | $<1\times10^{-14}$ | 1 | 0.57 | $<1\times10^{-14}$ | $<1\times10^{-14}$ | $<1\times10^{-14}$ | $9.31\times10^{-05}$ | $<1\times10^{-14}$ | $3.54\times10^{-10}$ | $<1\times10^{-14}$ | 0.038 | $7.07\times10^{-09}$ | $1.13\times10^{-11}$ |
| post-challenge glucose (mmol/l) | 0.94 | $<1\times10^{-14}$ | $<1\times10^{-14}$ | 0.046 | 0.4 | $<1\times10^{-14}$ | $<1\times10^{-14}$ | $<1\times10^{-14}$ | 0.087 | $<1\times10^{-14}$ | $3.63\times10^{-12}$ | $<1\times10^{-14}$ | $8.23\times10^{-07}$ | $<1\times10^{-14}$ | $2.19\times10^{-12}$ |
| glycated hemoglobin (mmol/mol) | 0.26 | $3.78\times10^{-09}$ | $6.54\times10^{-04}$ | 1 | 0.42 | $4.09\times10^{-08}$ | $2.52\times10^{-12}$ | $9.33\times10^{-07}$ | 0.43 | $2.84\times10^{-11}$ | $2.06\times10^{-06}$ | 0.044 | 0.71 | $1.57\times10^{-05}$ | 0.019 |
| triglycerides (mmol/l) | $1.98\times10^{-04}$ | 0.0032 | $1.47\times10^{-13}$ | 0.88 | 0.019 | $5.78\times10^{-05}$ | $1.15\times10^{-12}$ | $<1\times10^{-14}$ | $3.55\times10^{-04}$ | $<1\times10^{-14}$ | 0.0024 | $<1\times10^{-14}$ | 1 | $3.36\times10^{-05}$ | $9.77\times10^{-05}$ |
| insulin sensitivity (Matsuda) (AU) | $<1\times10^{-14}$ | $2.43\times10^{-04}$ | $<1\times10^{-14}$ | $1.13\times10^{-04}$ | $<1\times10^{-14}$ | $<1\times10^{-14}$ | $<1\times10^{-14}$ | $<1\times10^{-14}$ | $6.41\times10^{-10}$ | $<1\times10^{-14}$ | $<1\times10^{-14}$ | $<1\times10^{-14}$ | $7.87\times10^{-08}$ | $<1\times10^{-14}$ | 0.44 |
| fasting insulin (pmol/l) | $6.49\times10^{-06}$ | 0.98 | $4.08\times10^{-07}$ | 0.95 | $7.00\times10^{-04}$ | 0.62 | $<1\times10^{-14}$ | $<1\times10^{-14}$ | $<1\times10^{-14}$ | $<1\times10^{-14}$ | $<1\times10^{-14}$ | $<1\times10^{-14}$ | $<1\times10^{-14}$ | $<1\times10^{-14}$ | 0.02 |
| insulinogenic index (AU) | $3.96\times10^{-05}$ | $4.55\times10^{-09}$ | 0.55 | 0.52 | 0.044 | $9.95\times10^{-05}$ | 0.05 | 0.86 | 0.1 | 0.76 | 1 | $4.03\times10^{-06}$ | $2.21\times10^{-10}$ | 0.27 | 0.016 |
| disposition index (AU) | 0.97 | $3.65\times10^{-06}$ | $2.82\times10^{-04}$ | 0.96 | 1 | $3.31\times10^{-04}$ | $6.57\times10^{-05}$ | 0.0018 | 1 | 0.002 | $9.13\times10^{-04}$ | 0.026 | 0.71 | 0.029 | 0.76 |
| cholesterol (mmol/l) | 1 | 0.011 | 0.0058 | 0.95 | 0.99 | $7.25\times10^{-04}$ | 0.0073 | 0.0039 | 0.99 | $6.06\times10^{-04}$ | 0.24 | 0.15 | 0.8 | 0.033 | 0.55 |
| C-reactive protein (mg/dl) | 0.38 | 1 | 0.25 | 0.17 | $4.65\times10^{-04}$ | 0.37 | $2.96\times10^{-09}$ | $4.84\times10^{-13}$ | $4.98\times10^{-08}$ | $2.48\times10^{-04}$ | $3.32\times10^{-06}$ | $6.39\times10^{-11}$ | $5.14\times10^{-05}$ | 0.082 | 0.25 |
| LDL (mmol/l) | 0.011 | 0.33 | $4.18\times10^{-06}$ | 0.98 | 0.1 | 0.092 | 0.0072 | $1.29\times10^{-08}$ | 0.56 | 0.0011 | 0.79 | $5.77\times10^{-05}$ | 0.97 | 0.37 | 0.15 |
| HDL (mmol/l) | $<1\times10^{-14}$ | 1 | $<1\times10^{-14}$ | 0.37 | $<1\times10^{-14}$ | 0.74 | $4.94\times10^{-04}$ | $<1\times10^{-14}$ | 0.0045 | 0.15 | 0.51 | $<1\times10^{-14}$ | 0.87 | 1 | 0.065 |
| aspartate-aminotransferase (U/l) | 1 | 1 | 1 | 1 | 0.99 | 1 | $3.38\times10^{-14}$ | $3.75\times10^{-14}$ | $3.12\times10^{-14}$ | $3.44\times10^{-14}$ | 0.036 | 0.14 | 0.075 | 0.023 | $7.41\times10^{-10}$ |
| alanine-aminotransferase (U/l) | 0.77 | 1 | 0.66 | 0.98 | 0.36 | 1 | $<1\times10^{-14}$ | $<1\times10^{-14}$ | $<1\times10^{-14}$ | $<1\times10^{-14}$ | $7.76\times10^{-06}$ | $1.08\times10^{-08}$ | $7.30\times10^{-05}$ | $5.34\times10^{-04}$ | $3.99\times10^{-09}$ |
| gamma-glutamyl transferase (U/l) | 0.46 | 0.23 | 0.0015 | 1 | 0.81 | 0.085 | $1.64\times10^{-07}$ | $2.33\times10^{-11}$ | 0.0028 | $2.57\times10^{-08}$ | $7.62\times10^{-05}$ | $7.87\times10^{-09}$ | 0.27 | $1.18\times10^{-05}$ | 0.33 |
| serum creatinine (mg/dl) | 0.95 | 1 | 1 | 1 | 1 | 1 | 0.26 | 0.75 | 0.5 | 0.52 | 0.36 | 0.91 | 0.68 | 0.7 | 0.99 |
| urinary albumin-creatinine ratio (mg/g) | 1 | 1 | 0.99 | 1 | 1 | 1 | 0.6 | 0.78 | 0.44 | 0.66 | 1 | 0.99 | 1 | 1 | 0.47 |
| carotid intima media thickness | 1 | $1.61\times10^{-07}$ | $1.42\times10^{-05}$ | 0.95 | 0.99 | $6.99\times10^{-06}$ | $1.36\times10^{-06}$ | $3.11\times10^{-05}$ | 1 | $2.99\times10^{-05}$ | $4.06\times10^{-05}$ | 0.0016 | 0.6 | 0.0013 | 0.44 |
| polygenic risk score | 0.2 | 0.022 | 0.96 | 0.97 | 0.034 | 0.0022 | 0.59 | 1 | 0.9 | 0.22 | 1 | 0.28 | 0.038 | 0.92 | 0.7 |

#### Supplementary Table 2

Cohort characteristics of the **Whitehall II** cohort after stratification using the cluster medians established in the TUEF/TULIP cohort (n=6810). P-values were computed with one-way ANOVA for continuous variables and two-sided chi-squared tests for categorical variables.

| | 1 | 2 | 3 | 4 | 5 | 6 | p |
|---|---|---|---|---|---|---|---|
| **n** | 1362 | 4228 | 322 | 509 | 123 | 266 | |
| **sex = male (%)** | 1052 (77.2) | 2979 (70.5) | 220 (68.3) | 355 (69.7) | 76 (61.8) | 178 (66.9) | $1.57\times10^{-06}$ |
| **BMI (kg/m$^2$, mean (SD))** | 25.96 (2.03) | 23.72 (2.50) | 27.39 (2.58) | 30.17 (2.57) | 31.58 (3.68) | 32.84 (3.68) | $<2.23\times10^{-308}$ |
| **age (years, mean (SD))** | 50.96 (6.51) | 50.63 (6.73) | 53.73 (6.68) | 51.40 (6.40) | 54.15 (7.65) | 52.16 (7.12) | $1.52\times10^{-20}$ |
| **Waist circumference (cm, mean (SD))** | 89.43 (7.46) | 81.95 (9.60) | 92.62 (8.56) | 100.15 (7.61) | 103.25 (9.67) | 106.05 (8.78) | $<2.23\times10^{-308}$ |
| **Hip circumference (cm, mean (SD))** | 97.89 (4.48) | 94.90 (5.60) | 100.75 (5.63) | 107.00 (5.41) | 106.19 (6.77) | 109.69 (7.03) | $<2.23\times10^{-308}$ |
| **Systolic blood pressure (mmHg) (mean (SD))** | 122.99 (13.25) | 118.39 (13.25) | 129.51 (14.90) | 123.67 (13.00) | 132.59 (12.86) | 129.23 (14.18) | $2.94\times10^{-109}$ |
| **Diastolic blood pressure (mmHg) (mean (SD))** | 81.35 (9.06) | 77.17 (9.32) | 83.73 (10.51) | 81.86 (9.55) | 85.16 (9.51) | 85.04 (9.43) | $3.98\times10^{-110}$ |
| **Glucose (fasting, mmol/l) (mean (SD))** | 5.28 (0.42) | 5.13 (0.45) | 5.59 (0.53) | 5.19 (0.41) | 5.61 (0.54) | 5.39 (0.49) | $7.49\times10^{-109}$ |
| **Glucose (post-challenge, mmol/l) (mean (SD))** | 5.74 (1.16) | 5.17 (1.40) | 8.56 (1.17) | 5.01 (1.11) | 8.33 (1.41) | 5.86 (1.22) | $<2.23\times10^{-308}$ |
| **AUC glucose (mmol 2h l-1) (mean (SD))** | 661.52 (70.94) | 618.03 (90.47) | 848.98 (67.83) | 611.81 (69.17) | 836.33 (90.90) | 675.38 (73.66) | $<2.23\times10^{-308}$ |
| **Insulin (fasting, pmol/l) (mean (SD))** | 62.90 (28.16) | 30.81 (15.91) | 58.77 (26.53) | 50.99 (20.49) | 119.60 (41.87) | 122.35 (39.48) | $<2.23\times10^{-308}$ |
| **Insulin (post-challenge, pmol/l) (mean (SD))** | 441.42 (236.65) | 219.27 (153.28) | 595.78 (268.04) | 248.24 (148.82) | 884.65 (411.76) | 583.18 (288.61) | $<2.23\times10^{-308}$ |
| **Insulin secretion (Stumvoll) (AU, mean (SD))** | 1055.35 (319.60) | 752.75 (236.45) | 729.55 (348.63) | 915.20 (239.87) | 1422.77 (504.31) | 1528.23 (397.45) | $<2.23\times10^{-308}$ |
| **insulin sensitivity (Matsuda) (AU, mean (SD))** | 17.25 (5.80) | 41.43 (21.41) | 13.36 (4.74) | 26.17 (9.09) | 7.50 (2.67) | 9.76 (2.81) | $<2.23\times10^{-308}$ |
| **Triglycerides (mmol/l) (mean (SD))** | 1.88 (0.93) | 1.08 (0.53) | 1.78 (0.81) | 1.68 (0.84) | 2.93 (1.22) | 2.00 (0.92) | $<2.23\times10^{-308}$ |
| **HDL chol (mean (SD))** | 1.18 (0.27) | 1.60 (0.40) | 1.33 (0.33) | 1.18 (0.28) | 1.05 (0.26) | 1.23 (0.33) | $<2.23\times10^{-308}$ |
| **glycaemic category (%)** | | | | | | | $<2.23\times10^{-308}$ |
| **NGT** | 968 (71.1) | 3393 (80.3) | 24 ( 7.5) | 411 (80.7) | 20 (16.3) | 159 (59.8) | |
| **IFG** | 333 (24.4) | 617 (14.6) | 66 (20.5) | 93 (18.3) | 24 (19.5) | 94 (35.3) | |
| **IGT** | 56 ( 4.1) | 183 ( 4.3) | 141 (43.8) | 5 ( 1.0) | 42 (34.1) | 10 ( 3.8) | |
| **IFG+IGT** | 5 ( 0.4) | 35 ( 0.8) | 91 (28.3) | 0 ( 0.0) | 37 (30.1) | 3 ( 1.1) | |
| **Smoking (%)** | | | | | | | $1.50\times10^{-04}$ |
| **never** | 580 (42.6) | 1909 (45.2) | 143 (44.4) | 184 (36.1) | 60 (48.8) | 105 (39.5) | |
| **ex** | 538 (39.5) | 1567 (37.1) | 145 (45.0) | 213 (41.8) | 46 (37.4) | 119 (44.7) | |
| **current** | 177 (13.0) | 534 (12.6) | 20 ( 6.2) | 86 (16.9) | 11 ( 8.9) | 34 (12.8) | |
| **missing** | 67 ( 4.9) | 218 ( 5.2) | 14 ( 4.3) | 26 ( 5.1) | 6 ( 4.9) | 8 ( 3.0) | |
| **Antihypertensive medication = yes (%)** | 113 ( 8.3) | 199 ( 4.7) | 49 (15.2) | 51 (10.0) | 29 (23.6) | 45 (16.9) | $3.02\times10^{-34}$ |
| **Lipid lowering medication = yes (%)** | 9 ( 0.7) | 28 ( 0.7) | 6 ( 1.9) | 6 ( 1.2) | 5 ( 4.1) | 8 ( 3.0) | $2.37\times10^{-06}$ |

#### Supplementary Table 3

Post-hoc tests for comparison of cluster means in the **Whitehall II** study. Numbers denote p-values for all pairwise comparisons computed with Tukey's test (n=6810).

| | 2-1 | 3-1 | 3-2 | 4-1 | 4-2 | 4-3 | 5-1 | 5-2 | 5-3 | 5-4 | 6-1 | 6-2 | 6-3 | 6-4 | 6-5 |
|---|---|---|---|---|---|---|---|---|---|---|---|---|---|---|---|
| **age** | 0.64 | $3.48\times10^{-10}$ | $5.79\times10^{-13}$ | 0.79 | 0.14 | $1.55\times10^{-05}$ | $5.98\times10^{-06}$ | $1.43\times10^{-07}$ | 0.99 | $6.30\times10^{-04}$ | 0.078 | 0.0043 | 0.052 | 0.67 | 0.07 |
| **BMI** | $4.79\times10^{-13}$ | $5.43\times10^{-13}$ | $4.79\times10^{-13}$ | $4.79\times10^{-13}$ | $4.79\times10^{-13}$ | $4.79\times10^{-13}$ | $4.79\times10^{-13}$ | $4.79\times10^{-13}$ | $4.79\times10^{-13}$ | $2.99\times10^{-07}$ | $4.79\times10^{-13}$ | $4.79\times10^{-13}$ | $4.79\times10^{-13}$ | $4.79\times10^{-13}$ | $6.48\times10^{-05}$ |
| **Hip circumference** | $4.79\times10^{-13}$ | $5.38\times10^{-13}$ | $4.79\times10^{-13}$ | $4.79\times10^{-13}$ | $4.79\times10^{-13}$ | $4.79\times10^{-13}$ | $4.79\times10^{-13}$ | $4.79\times10^{-13}$ | $5.35\times10^{-13}$ | 0.68 | $4.79\times10^{-13}$ | $4.79\times10^{-13}$ | $4.79\times10^{-13}$ | $1.34\times10^{-09}$ | $7.28\times10^{-08}$ |
| **Waist circumference** | $4.79\times10^{-13}$ | $1.61\times10^{-07}$ | $4.79\times10^{-13}$ | $4.79\times10^{-13}$ | $4.79\times10^{-13}$ | $4.79\times10^{-13}$ | $4.79\times10^{-13}$ | $4.79\times10^{-13}$ | $5.01\times10^{-13}$ | 0.008 | $4.79\times10^{-13}$ | $4.79\times10^{-13}$ | $4.79\times10^{-13}$ | $5.29\times10^{-13}$ | 0.048 |
| **Systolic blood pressure (mmHg)** | $<1\times10^{-14}$ | $<1\times10^{-14}$ | $<1\times10^{-14}$ | 0.92 | $<1\times10^{-14}$ | $1.36\times10^{-08}$ | $<1\times10^{-14}$ | $<1\times10^{-14}$ | 0.25 | $4.56\times10^{-10}$ | $4.83\times10^{-11}$ | $<1\times10^{-14}$ | 1 | $5.59\times10^{-07}$ | 0.19 |
| **Diastolic blood pressure (mmHg)** | $1.55\times10^{-12}$ | $6.05\times10^{-04}$ | $1.55\times10^{-12}$ | 0.9 | $1.61\times10^{-12}$ | 0.057 | $2.14\times10^{-04}$ | $1.61\times10^{-12}$ | 0.7 | 0.0058 | $6.09\times10^{-08}$ | $1.55\times10^{-12}$ | 0.54 | $1.03\times10^{-04}$ | 1 |
| **Glucose (fasting, mmol/l)** | $4.79\times10^{-13}$ | $4.79\times10^{-13}$ | $4.79\times10^{-13}$ | $4.80\times10^{-13}$ | 0.61 | $4.79\times10^{-13}$ | $4.79\times10^{-13}$ | $4.79\times10^{-13}$ | 0.71 | $4.79\times10^{-13}$ | 0.13 | $5.35\times10^{-13}$ | $4.79\times10^{-13}$ | $5.35\times10^{-13}$ | $4.79\times10^{-13}$ |
| **Glucose (post-challenge, mmol/l)** | $5.34\times10^{-13}$ | $5.28\times10^{-13}$ | $4.79\times10^{-13}$ | $3.77\times10^{-04}$ | 0.11 | $4.79\times10^{-13}$ | $9.52\times10^{-13}$ | $4.79\times10^{-13}$ | 1 | $5.40\times10^{-13}$ | 0.0045 | $5.48\times10^{-13}$ | $2.00\times10^{-06}$ | $2.02\times10^{-08}$ | $1.82\times10^{-04}$ |
| **Insulin (fasting, pmol/l)** | $4.79\times10^{-13}$ | $4.79\times10^{-13}$ | $4.79\times10^{-13}$ | $5.35\times10^{-13}$ | 0.11 | $4.79\times10^{-13}$ | $4.79\times10^{-13}$ | $4.79\times10^{-13}$ | 0.57 | $4.79\times10^{-13}$ | 0.73 | $5.42\times10^{-13}$ | $4.79\times10^{-13}$ | $5.33\times10^{-13}$ | $4.79\times10^{-13}$ |
| **insulin sensitivity (Matsuda)** | $4.79\times10^{-13}$ | 0.027 | $4.79\times10^{-13}$ | $5.36\times10^{-13}$ | $4.79\times10^{-13}$ | $7.61\times10^{-06}$ | $4.79\times10^{-13}$ | $4.79\times10^{-13}$ | $4.79\times10^{-13}$ | $4.79\times10^{-13}$ | $4.79\times10^{-13}$ | $4.79\times10^{-13}$ | $4.79\times10^{-13}$ | $4.79\times10^{-13}$ | 0.86 |
| **Insulin (post-challenge, pmol/l)** | $4.79\times10^{-13}$ | 0.004 | $4.79\times10^{-13}$ | $5.34\times10^{-13}$ | $4.79\times10^{-13}$ | $5.36\times10^{-13}$ | $3.45\times10^{-08}$ | $4.79\times10^{-13}$ | 0.018 | $5.36\times10^{-13}$ | $1.70\times10^{-09}$ | $4.79\times10^{-13}$ | 0.12 | $4.79\times10^{-13}$ | 0.84 |
| **Insulin secretion (Stumvoll)** | $4.79\times10^{-13}$ | $4.79\times10^{-13}$ | $4.79\times10^{-13}$ | $4.79\times10^{-13}$ | 0.017 | $4.79\times10^{-13}$ | $4.79\times10^{-13}$ | $4.79\times10^{-13}$ | $4.79\times10^{-13}$ | $4.79\times10^{-13}$ | $5.24\times10^{-13}$ | $4.79\times10^{-13}$ | 0.97 | $4.79\times10^{-13}$ | $4.79\times10^{-13}$ |
| **Triglycerides (mmol/l)** | $4.79\times10^{-13}$ | $4.79\times10^{-13}$ | 0.69 | $5.41\times10^{-13}$ | $4.79\times10^{-13}$ | $5.32\times10^{-13}$ | $4.79\times10^{-13}$ | $4.79\times10^{-13}$ | $4.79\times10^{-13}$ | $4.79\times10^{-13}$ | $4.79\times10^{-13}$ | $4.79\times10^{-13}$ | $4.79\times10^{-13}$ | $4.79\times10^{-13}$ | 0.006 |
| **HDL cholesterol** | $4.79\times10^{-13}$ | 0.23 | $4.79\times10^{-13}$ | $5.02\times10^{-07}$ | $4.79\times10^{-13}$ | 0.29 | $4.79\times10^{-13}$ | $4.79\times10^{-13}$ | $4.79\times10^{-13}$ | $4.79\times10^{-13}$ | 0.12 | $4.79\times10^{-13}$ | 0.0029 | $2.45\times10^{-08}$ | $4.79\times10^{-13}$ |

#### Supplementary Table 4

Association of clusters with genotype for every tested variant. Tested top diabetes-related variants that were genotyped in TUEF/TULIP were taken from Mahajan et al 2018. Results show one-way ANOVA for each variant between cluster as factor variable and genotype as continuous outcome. P-value adjustment for multiple testing was performed with Benjamini-Hochberg correction.

| | | F | P-VALUE | ADJUSTED P |
|---|---|---|---|---|
| **1** | MTNR1B_rs10830963_G | 4.806 | 0.00024 | 0.02 |
| **2** | TCF7L2_rs7903146_T | 3.442 | 0.004 | 0.1 |
| **3** | KCNQ1_rs2237892_T | 2.953 | 0.01 | 0.2 |
| **4** | PNPLA3_rs738409_C | 2.355 | 0.04 | 0.6 |
| **5** | TM6SF2_rs58542926_T | 2.281 | 0.04 | 0.6 |
| **6** | CDKAL1_rs7756992_G | 2.016 | 0.07 | 0.7 |
| **7** | JAZF1_rs864745_G | 1.987 | 0.08 | 0.7 |
| **8** | HNF1B_rs7501939_T | 1.909 | 0.09 | 0.7 |
| **9** | VEGFA_rs6905288_G | 1.884 | 0.09 | 0.7 |
| **10** | TFAP2B_rs2206277_A | 1.69 | 0.1 | 0.8 |
| **11** | ZMIZ1_rs12571751_G | 1.669 | 0.1 | 0.8 |
| **12** | BCL11A_rs243021_T | 1.558 | 0.2 | 0.8 |
| **13** | C2CD4A-B_rs4502156_C | 1.545 | 0.2 | 0.8 |
| **14** | HNF1A_rs1169288_G | 1.501 | 0.2 | 0.8 |
| **15** | SLC30A8_rs13266634_T | 1.498 | 0.2 | 0.8 |
| **16** | WFS1_rs4689388_G | 1.439 | 0.2 | 0.8 |
| **17** | GIPR_GSA-rs8108269_G | 1.35 | 0.2 | 0.8 |
| **18** | ABO_exm-rs505922_C | 1.317 | 0.3 | 0.8 |
| **19** | CENPW_rs1361108_T | 1.253 | 0.3 | 0.8 |
| **20** | IRS1_rs2943641_T | 1.179 | 0.3 | 0.8 |
| **21** | ANK1_rs516946_A | 1.178 | 0.3 | 0.8 |
| **22** | MC4R_rs17782313_C | 1.174 | 0.3 | 0.8 |
| **23** | DGKB_rs2191349_G | 1.114 | 0.4 | 0.8 |
| **24** | PRC1_rs8042680_A | 1.109 | 0.4 | 0.8 |
| **25** | PAM_GSA-rs35658696_G | 1.108 | 0.4 | 0.8 |
| **26** | TMEM258_rs102275_G | 1.087 | 0.4 | 0.8 |
| **27** | HMG20A_GSA-rs7177055_G | 1.053 | 0.4 | 0.8 |
| **28** | NFAT5_rs1364063_C | 1.049 | 0.4 | 0.8 |
| **29** | FTO_rs1558902_A | 1.046 | 0.4 | 0.8 |
| **30** | PAX4_rs2233580_A | 0.837 | 0.5 | 1 |
| **31** | RREB1_rs9379084_A | 0.83 | 0.5 | 1 |
| **32** | POC5_rs2307111_G | 0.82 | 0.5 | 1 |
| **33** | CENTD2_rs11603334_A | 0.804 | 0.5 | 1 |
| **34** | CCND2_rs11063069_G | 0.774 | 0.6 | 1 |
| **35** | GIPR_rs1800437_C | 0.767 | 0.6 | 1 |
| **36** | CEP68_rs7572857_A | 0.749 | 0.6 | 1 |
| **37** | GRB14_rs13389219_T | 0.74 | 0.6 | 1 |
| **38** | CMIP_rs2925979_A | 0.725 | 0.6 | 1 |
| **39** | HNF4A_rs1800961_T | 0.718 | 0.6 | 1 |
| **40** | ZBED3_rs4457053_G | 0.701 | 0.6 | 1 |
| **41** | MRAS_rs2306374_C | 0.668 | 0.6 | 1 |
| **42** | ARL15_rs6450176_A | 0.66 | 0.7 | 1 |

| | | | | |
|---|---|---|---|---|
| **43** | PPARG_rs1801282_G | 0.649 | 0.7 | 1 |
| **44** | TOMM40-APOE_rs769449_A | 0.595 | 0.7 | 1 |
| **45** | ZHX3_rs17265513_C | 0.565 | 0.7 | 1 |
| **46** | MPHOSPH9_GSA-rs1727307_A | 0.529 | 0.8 | 1 |
| **47** | BCAR1_rs7202877_G | 0.526 | 0.8 | 1 |
| **48** | ADAMTS9_rs4607103_T | 0.511 | 0.8 | 1 |
| **49** | CDKN2A-B_rs10965250_A | 0.494 | 0.8 | 1 |
| **50** | NRXN3_rs10146997_G | 0.485 | 0.8 | 1 |
| **51** | KLF14_rs972283_A | 0.431 | 0.8 | 1 |
| **52** | KCNQ1_rs2237895_C | 0.398 | 0.9 | 1 |
| **53** | PEPD_rs731839_C | 0.388 | 0.9 | 1 |
| **54** | HHEX-IDE_rs5015480_T | 0.357 | 0.9 | 1 |
| **55** | SPRY2_rs1359790_T | 0.312 | 0.9 | 1 |
| **56** | HMGA2_rs1531343_C | 0.272 | 0.9 | 1 |
| **57** | PROX1_rs340874_A | 0.268 | 0.9 | 1 |
| **58** | ADCY5_rs11708067_G | 0.253 | 0.9 | 1 |
| **59** | CDC123/CAMK1D_rs10906115_G | 0.197 | 1 | 1 |
| **60** | HNF1A_GSA-rs1800574_T | 0.193 | 1 | 1 |
| **61** | ANKRD55_rs459193_T | 0.173 | 1 | 1 |
| **62** | KCNJ11_rs5219_T | 0.063 | 1 | 1 |
| **63** | TSPAN8_rs7961581_C | 0.043 | 1 | 1 |

#### Supplementary Table 5

Cluster-wise hazard ratios for different outcomes in the Whitehall II cohort. Clusters and the denoted adjustment covariates were applied in proportional hazards models. Values show point estimates as hazard ratios with 95% confidence intervals.

| outcome | adjustment | n | cluster2 | cluster3 | cluster4 | cluster5 | cluster6 |
|---|---|---|---|---|---|---|---|
| **diabetes** | | 6643 | 0.40*** (0.33-0.47) | 3.45*** (2.76-4.31) | 0.80 (0.61-1.06) | 6.62*** (5.06-8.67) | 2.22*** (1.70-2.89) |
| **diabetes** | sex, age, age2, BMI | 6643 | 0.45*** (0.38-0.55) | 3.02*** (2.41-3.79) | 0.67* (0.50-0.90) | 5.07*** (3.77-6.82) | 1.66** (1.21-2.27) |
| **diabetes** | sex, age, age2, BMI, baseline glucose AUC | 6643 | 0.56*** (0.46-0.67) | 1.50** (1.15-1.95) | 0.79 (0.58-1.07) | 2.67*** (1.93-3.69) | 1.49* (1.09-2.05) |
| **renal (CKD)** | | 5182 | 1.11 (0.96-1.27) | 1.47** (1.13-1.91) | 1.11 (0.88-1.40) | 1.60* (1.07-2.37) | 1.63*** (1.25-2.12) |
| **renal (CKD)** | sex, age, age2, BMI | 5182 | 1.03 (0.88-1.19) | 1.49** (1.14-1.94) | 1.14 (0.89-1.46) | 1.61* (1.06-2.43) | 1.61** (1.20-2.15) |
| **renal (CKD)** | sex, age, age2, BMI, fasting glucose, systolic BP, diastolic BP | 5174 | 1.01 (0.87-1.18) | 1.45* (1.11-1.90) | 1.13 (0.88-1.45) | 1.55* (1.02-2.35) | 1.63** (1.22-2.19) |
| **coronary heart disease** | | 6537 | 0.64*** (0.54-0.76) | 1.38* (1.03-1.86) | 1.24 (0.96-1.61) | 1.65* (1.08-2.53) | 1.22 (0.87-1.70) |
| **coronary heart disease** | sex, age, age2, BMI | 6537 | 0.72*** (0.60-0.87) | 1.19 (0.88-1.61) | 1.13 (0.85-1.49) | 1.38 (0.88-2.15) | 1.05 (0.72-1.52) |
| **coronary heart disease** | sex, age, age2, BMI, fasting glucose, systolic BP, diastolic BP | 6523 | 0.73** (0.61-0.88) | 1.21 (0.89-1.64) | 1.12 (0.85-1.48) | 1.40 (0.89-2.20) | 1.04 (0.72-1.52) |
| **CHD + stroke incidence** | | 6704 | 0.64*** (0.55-0.75) | 1.12 (0.85-1.47) | 1.25* (1.00-1.56) | 1.52* (1.05-2.19) | 1.03 (0.76-1.40) |
| **CHD + stroke incidence** | sex, age, age2, BMI | 6704 | 0.70*** (0.60-0.83) | 0.97 (0.74-1.28) | 1.16 (0.91-1.48) | 1.27 (0.86-1.87) | 0.89 (0.64-1.25) |
| **CHD + stroke incidence** | sex, age, age2, BMI, fasting glucose, systolic BP, diastolic BP | 6690 | 0.73*** (0.62-0.85) | 0.99 (0.75-1.31) | 1.17 (0.92-1.49) | 1.31 (0.88-1.94) | 0.90 (0.64-1.25) |
| **all-cause mortality** | | 6803 | 0.66*** (0.56-0.78) | 1.00 (0.73-1.38) | 0.84 (0.63-1.11) | 1.66* (1.11-2.48) | 1.47* (1.09-1.99) |
| **all-cause mortality** | sex, age, age2, BMI | 6803 | 0.68*** (0.57-0.81) | 0.80 (0.58-1.10) | 0.85 (0.63-1.15) | 1.43 (0.94-2.19) | 1.45* (1.03-2.05) |
| **all-cause mortality** | sex, age, age2, BMI, fasting glucose, systolic BP, diastolic BP | 6789 | 0.68*** (0.57-0.82) | 0.78 (0.56-1.09) | 0.85 (0.63-1.15) | 1.42 (0.93-2.18) | 1.45* (1.02-2.04) |

Legend: Hazard ratios with 95% confidence intervals compared to cluster 1. Corresponding p-values are shown with: *** for a p.value < 0.0005, ** for a p.value < 0.005 and * for a p.value <0.05. CKD: chronic kidney disease Stage 3 or worse (eGFR<60 ml/min/1.73m$^2$), CHD: coronary heart disease

#### Supplementary Table 6

Cox multivariable proportional hazards model for diabetes as outcome in Whitehall II. Estimate indicates adjusted hazard ratio (exp(β)) with 95% confidence interval, n=6643 individuals with follow-up.

| | **Diabetes** | | |
|---|---|---|---|
| *Predictors* | *Estimates* | *CI* | *p* |
| cluster [2] | 0.631 | 0.509 – 0.782 | $2.59\times10^{-05}$ |
| cluster [3] | 2.68 | 2.12 – 3.38 | $1.12\times10^{-16}$ |
| cluster [4] | 0.687 | 0.508 – 0.929 | 0.015 |
| cluster [5] | 2.99 | 2.16 – 4.13 | $3.72\times10^{-11}$ |
| cluster [6] | 1.4 | 1.01 – 1.94 | 0.043 |
| sex: female | 1.11 | 0.926 – 1.34 | 0.26 |
| age [1st degree] | 90108345 | 247024 – $3.29\times10^{10}$ | $1.16\times10^{-09}$ |
| age [2nd degree] | 57124 | 134 – 24355955 | $3.92\times10^{-04}$ |
| BMI | 1.05 | 1.02 – 1.07 | $6.22\times10^{-04}$ |
| Fasting glucose baseline | 2.23 | 1.92 – 2.6 | $3.87\times10^{-25}$ |
| Smoking [ex] | 1.14 | 0.975 – 1.33 | 0.1 |
| Smoking [current] | 2.01 | 1.64 – 2.46 | $1.25\times10^{-11}$ |
| Smoking [missing information] | 1.38 | 0.996 – 1.9 | 0.053 |
| Triglycerides baseline | 1.14 | 1.04 – 1.24 | 0.006 |
| Cholesterol_baseline | 1.04 | 0.976 – 1.12 | 0.21 |
| HDL baseline | 0.7 | 0.535 – 0.916 | 0.0093 |

Supplementary Table 7

Cluster transitions to Ahqlvist-diabetes classes (N=201)

| Pre-diabetic cluster | **Total eligible for Ahlqvist-classification** | **2/SIDD** | **3/SIRD** | **4/MOD** | **5/MARD** |
|---|---|---|---|---|---|
| **1** | 817 | 6 (12%) | 8 (17%) | 11 (23%) | 23 (48%) |
| **2** | 2552 | 2 (3%) | 1 (2%) | 15 (24%) | 44 (71%) |
| **3** | 191 | 5 (12%) | 4 (10%) | 13 (31%) | 20 (48%) |
| **4** | 314 | 0 (0%) | 2 (14%) | 8 (57%) | 4 (29%) |
| **5** | 65 | 2 (17%) | 4 (33%) | 2 (17%) | 4 (33%) |
| **6** | 153 | 2 (9%) | 13 (57%) | 8 (35%) | 0 (0%) |

#### Supplementary Table 8

Cox multivariable proportional hazards model for stage 3 chronic kidney disease as outcome in Whitehall II. Estimate indicates adjusted hazard ratio (exp(β)) with 95% confidence interval and two-sided p-value, n=5126 individuals with follow-up.

| | **CKD3 or worse** | | |
|---|---|---|---|
| *Predictors* | *Estimates* | *CI* | *p* |
| cluster [2] | 0.929 | 0.781 – 1.1 | 0.4 |
| cluster [3] | 1.37 | 1.05 – 1.8 | 0.022 |
| cluster [4] | 1.16 | 0.898 – 1.49 | 0.26 |
| cluster [5] | 1.74 | 1.11 – 2.72 | 0.015 |
| cluster [6] | 1.63 | 1.2 – 2.22 | 0.0016 |
| age [1st degree] | 5.57E-10 | $1.19\times10^{-11}$ – $2.6\times10^{-8}$ | $1.65\times10^{-27}$ |
| age [2nd degree] | 7.90E+07 | 2259923 – $2.76\times10^{9}$ | $1.15\times10^{-23}$ |
| sex: female | 1.19 | 1.03 – 1.36 | 0.017 |
| Smoking [ex] | 1.1 | 0.985 – 1.23 | 0.091 |
| Smoking [current] | 1.47 | 1.14 – 1.9 | 0.0034 |
| Smoking [missing] | 0.666 | 0.494 – 0.897 | 0.0076 |
| BMI | 0.996 | 0.974 – 1.02 | 0.68 |
| Systolic blood pressure | 1.03 | 1.02 – 1.03 | $1.67\times10^{-17}$ |
| Diastolic blood pressure | 0.969 | 0.961 – 0.977 | $8.56\times10^{-13}$ |
| Glucose baseline | 0.887 | 0.784 – 1 | 0.057 |
| Triglycerides baseline | 0.581 | 0.000818 – 412 | 0.87 |
| Cholesterol baseline | 4.32 | $2.31\times10^{-6}$ – 8058826 | 0.84 |
| HDL baseline | 0.342 | $1.84\times10^{-7}$ – 637599 | 0.88 |
| LDL baseline | 0.219 | $1.18\times10^{-7}$ – 409504 | 0.84 |

#### Supplementary Table 9

Differences in renal sinus fat (RSF, mean of left and right) levels among clusters. Statistical analysis was performed with one-way ANOVA adjusted for sex, age, age$^2$ (A) or sex, age, age$^2$ and visceral fat (C). RSF, visceral fat and BMI were log$_e$-transformed. Post-hoc test of pairwise differences between specific clusters was computed with Tukey's HSD test for both models (Table B and D).

Model 1 (n=519 individuals with RSF>0): RSF ~ sex + BMI + Age + Age$^2$ + cluster

Model 2 (n=519 individuals with RSF>0): RSF ~ sex + BMI + Age + Age$^2$ + visceral fat + cluster

##### A. Analysis of variance table for model 1

| | TERM | DF | SUMSQ | MEANSQ | STATISTIC | P.VALUE |
|---|---|---|---|---|---|---|
| **1** | SEX | 1 | 19.72 | 19.72 | 51.59 | $2.44\times10^{-12}$ |
| **2** | BMI | 1 | 55.34 | 55.34 | 144.76 | $1.62\times10^{-29}$ |
| **3** | poly(AGE, 2) | 2 | 84.82 | 42.41 | 110.92 | $1.04\times10^{-40}$ |
| **4** | cluster | 5 | 15.89 | 3.18 | 8.31 | $1.44\times10^{-07}$ |
| **5** | Residuals | 509 | 194.6 | 0.38 | | |

##### B. Pairwise comparisons of clusters (Tukey) for model 1

| | CLUSTER VS | CLUSTER | ESTIMATE | STD.ERROR | DF | STATISTIC | P.VALUE |
|---|---|---|---|---|---|---|---|
| **1** | 1 | 2 | -0.05 | 0.11 | 509 | -0.49 | 1 |
| **2** | 1 | 3 | -0.29 | 0.13 | 509 | -2.22 | 0.23 |
| **3** | 1 | 4 | -0.3 | 0.11 | 509 | -2.81 | 0.058 |
| **4** | 1 | 5 | -0.33 | 0.11 | 509 | -3 | 0.034 |
| **5** | 1 | 6 | -0.6 | 0.1 | 509 | -5.97 | $6.7\times10^{-08}$ |
| **6** | 2 | 3 | -0.23 | 0.13 | 509 | -1.74 | 0.5 |
| **7** | 2 | 4 | -0.25 | 0.12 | 509 | -2.07 | 0.3 |
| **8** | 2 | 5 | -0.28 | 0.13 | 509 | -2.23 | 0.23 |
| **9** | 2 | 6 | -0.55 | 0.12 | 509 | -4.66 | $5.8\times10^{-05}$ |
| **10** | 3 | 4 | -0.01 | 0.13 | 509 | -0.09 | 1 |
| **11** | 3 | 5 | -0.05 | 0.13 | 509 | -0.36 | 1 |
| **12** | 3 | 6 | -0.32 | 0.12 | 509 | -2.63 | 0.093 |
| **13** | 4 | 5 | -0.03 | 0.1 | 509 | -0.33 | 1 |
| **14** | 4 | 6 | -0.3 | 0.09 | 509 | -3.36 | 0.011 |
| **15** | 5 | 6 | -0.27 | 0.08 | 509 | -3.36 | 0.011 |

##### C. Analysis of variance table for model 2

| | TERM | DF | SUMSQ | MEANSQ | STATISTIC | P.VALUE |
|---|---|---|---|---|---|---|
| **1** | SEX | 1 | 19.72 | 19.72 | 59.64 | $6.10\times10^{-14}$ |
| **2** | BMI | 1 | 55.34 | 55.34 | 167.35 | $2.75\times10^{-33}$ |
| **3** | poly(AGE, 2) | 2 | 84.82 | 42.41 | 128.23 | $8.25\times10^{-46}$ |
| **4** | VAT | 1 | 37.35 | 37.35 | 112.95 | $5.87\times10^{-24}$ |
| **5** | cluster | 5 | 5.14 | 1.03 | 3.11 | 0.009 |
| **6** | Residuals | 508 | 168 | 0.33 | | |

#### D. Pairwise comparisons of clusters (Tukey) for model 2

| | CLUSTER VS | CLUSTER | ESTIMATE | STD.ERROR | DF | STATISTIC | P.VALUE |
|---|---|---|---|---|---|---|---|
| **1** | 1 | 2 | -0.14 | 0.1 | 508 | -1.38 | 0.7 |
| **2** | 1 | 3 | -0.05 | 0.12 | 508 | -0.42 | 1 |
| **3** | 1 | 4 | -0.07 | 0.1 | 508 | -0.65 | 1 |
| **4** | 1 | 5 | 0.09 | 0.11 | 508 | 0.82 | 1 |
| **5** | 1 | 6 | -0.18 | 0.11 | 508 | -1.7 | 0.5 |
| **6** | 2 | 3 | 0.09 | 0.13 | 508 | 0.66 | 1 |
| **7** | 2 | 4 | 0.07 | 0.12 | 508 | 0.61 | 1 |
| **8** | 2 | 5 | 0.23 | 0.13 | 508 | 1.77 | 0.5 |
| **9** | 2 | 6 | -0.04 | 0.12 | 508 | -0.34 | 1 |
| **10** | 3 | 4 | -0.01 | 0.12 | 508 | -0.12 | 1 |
| **11** | 3 | 5 | 0.14 | 0.12 | 508 | 1.19 | 0.8 |
| **12** | 3 | 6 | -0.13 | 0.11 | 508 | -1.12 | 0.9 |
| **13** | 4 | 5 | 0.16 | 0.1 | 508 | 1.63 | 0.6 |
| **14** | 4 | 6 | -0.11 | 0.09 | 508 | -1.3 | 0.8 |
| **15** | 5 | 6 | -0.27 | 0.07 | 508 | -3.63 | 0.004 |

#### Supplementary Table 10

Cox multivariable proportional hazards model for all-cause mortality as outcome in Whitehall II. Estimate indicates adjusted hazard ratio (exp(β)) with 95% confidence interval and two-sided p-value, n=6789 individuals with follow-up.

| | Death from all causes | | |
|---|---|---|---|
| *Predictors* | *Estimates* | *CI* | *p* |
| cluster [2] | 0.8 | 0.651 – 0.983 | 0.034 |
| cluster [3] | 0.821 | 0.59 – 1.14 | 0.24 |
| cluster [4] | 0.823 | 0.608 – 1.11 | 0.21 |
| cluster [5] | 1.25 | 0.804 – 1.95 | 0.32 |
| cluster [6] | 1.45 | 1.02 – 2.08 | 0.039 |
| age [1st degree] | $1.67\times10^{26}$ | $1.89\times10^{23}$ – $1.47\times10^{29}$ | $3.27\times10^{-68}$ |
| age [2nd degree] | 5.41 | 0.0146 – 2010 | 0.58 |
| sex: female | 0.911 | 0.759 – 1.09 | 0.31 |
| BMI | 0.993 | 0.966 – 1.02 | 0.63 |
| Systolic blood pressure | 1 | 0.996 – 1.01 | 0.32 |
| Diastolic blood pressure | 1 | 0.994 – 1.02 | 0.4 |
| Fasting glucose baseline | 0.966 | 0.826 – 1.13 | 0.66 |
| Triglycerides baseline | 1.09 | 0.987 – 1.21 | 0.087 |
| Cholesterol baseline | 1.01 | 0.941 – 1.08 | 0.86 |
| HDL baseline | 0.883 | 0.692 – 1.13 | 0.32 |
| Smoking [ex] | 1.26 | 1.07 – 1.48 | 0.0047 |
| Smoking [current] | 2.51 | 2.07 – 3.04 | $1.35\times10^{-20}$ |
| Smoking [missing] | 1.28 | 0.9 – 1.82 | 0.17 |

#### Supplementary Table 11

Cohort characteristics of the TUEF/TULIP showing the full set and the subset the partitioning was performed on. Values denote means (SD).

| | **all** | **clustered** |
|---|---|---|
| n | 2771 | 899 |
| sex = male (%) | 1058 (38.2) | 346 (38.5) |
| age (mean (SD)) | 42.14 (14.35) | 44.61 (13.36) |
| BMI (kg/m2) (mean (SD)) | 27.95 (6.15) | 29.84 (5.73) |
| waist circumference (cm) (mean (SD)) | 92.85 (15.33) | 96.26 (14.76) |
| total adipose tissue MRI (liter) (mean (SD)) | 35.64 (13.93) | 35.95 (13.95) |
| subcutaneous adipose tissue MRI (liter) (mean (SD)) | 12.41 (6.42) | 12.44 (6.44) |
| visceral adipose tissue MRI (liter) (mean (SD)) | 3.73 (2.40) | 3.80 (2.42) |
| subcutaneous to visceral adipose ratio (mean (SD)) | 4.49 (2.85) | 4.35 (2.72) |
| visceral adipose % of total (mean (SD)) | 0.11 (0.06) | 0.11 (0.06) |
| liver fat content (mean (SD)) | 6.48 (6.78) | 7.06 (7.01) |
| fatty-liver disease (%) = yes (%) | 392 (36.9) | 341 (37.9) |
| renal sinus fat (mean of r&l, %) (mean (SD)) | 10.67 (6.37) | 10.76 (6.35) |
| systolic blood pressure (mmHg, mean (SD)) | 129.80 (18.31) | 131.45 (17.91) |
| diastolic blood pressure (mmHg, mean (SD)) | 82.69 (13.87) | 83.60 (12.27) |
| heart rate (BPM, mean (SD)) | 70.84 (10.92) | 70.08 (10.69) |
| fasting glucose (mmol/l) (mean (SD)) | 5.17 (0.56) | 5.35 (0.57) |
| post-challenge glucose @ 30 min (mmol/l, mean (SD)) | 8.44 (1.56) | 8.87 (1.54) |
| post-challenge glucose @ 60 min (mmol/l, mean (SD)) | 8.20 (2.40) | 8.95 (2.37) |
| post-challenge glucose @ 90 min (mmol/l, mean (SD)) | 6.95 (2.09) | 7.63 (2.14) |
| post-challenge glucose @ 120 min (mmol/l, mean (SD)) | 6.23 (1.61) | 6.73 (1.54) |
| glycaemic category (%) | | |
| NGT | 1960 (70.7) | 529 (58.8) |
| IFG | 360 (13.0) | 168 (18.7) |
| IGT | 247 ( 8.9) | 98 (10.9) |
| IFG+IGT | 204 ( 7.4) | 104 (11.6) |
| GAD antibody = TRUE (%) | 36 ( 4.7) | 17 ( 4.9) |
| triglycerides (mmol/l) (mean (SD)) | 1.37 (1.40) | 1.37 (0.86) |
| insulin sensitivity (Matsuda) (AU, mean (SD)) | 18.87 (12.67) | 13.95 (8.66) |
| fasting insulin (pmol/l) (mean (SD)) | 53.39 (38.72) | 64.46 (44.40) |
| insulinogenic index (AU, mean (SD)) | 128.26 (151.49) | 141.55 (165.61) |
| disposition index (AU, mean (SD)) | 2106.75 (3773.09) | 1823.42 (3688.84) |
| C-reactive protein (mg/dl, mean (SD)) | 0.27 (0.36) | 0.27 (0.38) |
| LDL cholesterol (mmol/l) (mean (SD)) | 3.04 (0.87) | 3.06 (0.85) |
| HDL cholesterol (mmol/l) (mean (SD)) | 1.42 (0.37) | 1.36 (0.34) |
| aspartate-aminotransferase (U/l, mean (SD)) | 22.25 (9.47) | 24.06 (9.33) |
| alanine-aminotransferase (U/l, mean (SD)) | 27.71 (17.34) | 29.09 (18.83) |
| gamma-glutamyl transferase (U/l, mean (SD)) | 25.39 (27.40) | 26.69 (23.74) |
| serum creatinine (mg/dl) (mean (SD)) | 0.82 (0.17) | 0.81 (0.17) |
| urinary albumin-creatinine ratio (mean (SD)) | 18.88 (47.13) | 17.84 (28.92) |
| polygenic risk score (mean (SD)) | -0.02 (1.01) | 0.01 (0.94) |
| family history of diabetes (%) | | |
| a_no family history | 360 (41.0) | 168 (39.5) |
| b_second degree relative | 185 (21.1) | 94 (22.1) |
| c_first degree relative | 333 (37.9) | 163 (38.4) |
| ever smoked = yes (%) | 411 (42.5) | 215 (48.0) |
| current smoking = yes (%) | 8 ( 1.1) | 5 ( 1.3) |
| cholesterol lowering medication = yes (%) | 66 (2.4) | 23 (2.6) |
| antihypertensive medication = yes (%) | 219 (7.9) | 105 (11.7) |

#### Supplementary Table 12

Cohort characteristics of the analysed set from the Whitehall-II cohort also showing what proportion of subjects was assigned to the clusters in specific study phases (upon first availability of the complete set of variables needed for cluster assignment). Values denote means (SD).

| | **Overall** |
|---|---|
| n | 6810 |
| sex = male (%) | 4860 (71.4) |
| cluster (%) | |
| 1 | 1362 (20.0) |
| 2 | 4228 (62.1) |
| 3 | 322 ( 4.7) |
| 4 | 509 ( 7.5) |
| 5 | 123 ( 1.8) |
| 6 | 266 ( 3.9) |
| clustered at (%) | |
| Phase03 | 5855 (86.0) |
| Phase05 | 478 ( 7.0) |
| Phase07 | 374 ( 5.5) |
| Phase09 | 103 ( 1.5) |
| BMI (kg/m$^2$, mean (SD)) | 25.32 (3.58) |
| age (years, mean (SD)) | 51.03 (6.74) |
| Waist circumference (cm, mean (SD)) | 86.64 (11.45) |
| Hip circumference (mean (SD)) | 97.46 (6.97) |
| Systolic blood pressure (mmHg) (mean (SD)) | 120.91 (13.86) |
| Diastolic blood pressure (mmHg) (mean (SD)) | 79.12 (9.71) |
| Glucose (fasting, mmol/l) (mean (SD)) | 5.21 (0.47) |
| Glucose (post-challenge, mmol/l) (mean (SD)) | 5.52 (1.55) |
| AUC glucose (mmol 2h l-1) (mean (SD)) | 643.37 (101.35) |
| Insulin (fasting, pmol/l) (mean (SD)) | 45.24 (31.74) |
| Insulin (post-challenge, pmol/l) (mean (SD)) | 309.90 (242.27) |
| Insulin secretion (Stumvoll) (AU, mean (SD)) | 866.71 (338.79) |
| insulin sensitivity (Matsuda) (AU, mean (SD)) | 32.27 (21.14) |
| Triglycerides (mmol/l) (mean (SD)) | 1.39 (0.82) |
| HDL cholesterol (mmol/l) (mean (SD)) | 1.45 (0.41) |
| glycaemic category (%) | |
| NGT | 4975 (73.1) |
| IFG | 1227 (18.0) |
| IGT | 437 ( 6.4) |
| IFG+IGT | 171 ( 2.5) |
| Smoking (%) | |
| never | 2981 (43.8) |
| ex | 2628 (38.6) |
| current | 862 (12.7) |
| missing | 339 ( 5.0) |
| Antihypertensive medication = yes (%) | 486 ( 7.1) |
| Lipid lowering medication = yes (%) | 62 ( 0.9) |

Supplementary Table 13

Correlation of clustering variables in the TUEF/TULIP cohort (n=899 individuals). The numbers indicate Spearman correlation coefficients.

| variables | AUC glu | genetic risk | HDL chol | insulin secr | insulin sensitivity | liver fat | sq adip | visc adip |
|---|---|---|---|---|---|---|---|---|
| AUC glu | 1 | 0.06 | -0.08 | -0.09 | -0.53 | 0.44 | 0.12 | 0.42 |
| genetic risk | 0.06 | 1 | -0.04 | -0.04 | 0.01 | 0.05 | -0.04 | -0.05 |
| HDL cholesterol | -0.08 | -0.04 | 1 | -0.26 | 0.28 | -0.19 | -0.27 | -0.26 |
| insulin secretion | -0.09 | -0.04 | -0.26 | 1 | -0.47 | 0.28 | 0.31 | 0.22 |
| insulin sensitivity | -0.53 | 0.01 | 0.28 | -0.47 | 1 | -0.48 | -0.36 | -0.52 |
| liver fat | 0.44 | 0.05 | -0.19 | 0.28 | -0.48 | 1 | 0.32 | 0.55 |
| sq adipose | 0.12 | -0.04 | -0.27 | 0.31 | -0.36 | 0.32 | 1 | 0.55 |
| visc adipose | 0.42 | -0.05 | -0.26 | 0.22 | -0.52 | 0.55 | 0.55 | 1 |

#### Supplementary Table 14

Sex-stratified cohort means in the **TUEF/TULIP** (n=899) and **Whitehall II** (n=6810) dataset for the proxy variables which were available in both cohorts. For variable units and dimensions, please refer to Supplementary Table 12.

| | | TUEF | | WHITEHALL-II | |
|---|---|---|---|---|---|
| | SEX | TUEF_mean | TUEF_sd | WH-II_mean | WH-II_sd |
| **AUC_GLUCOSE** | female | 722.01 | 107.8 | 643.48 | 103.39 |
| **BMI** | female | 29.66 | 5.98 | 25.51 | 4.58 |
| **HDL** | female | 1.46 | 0.35 | 1.72 | 0.43 |
| **HIP** | female | 109.07 | 12.87 | 97.73 | 9.23 |
| **INSU_F** | female | 9.22 | 6.26 | 6.16 | 4.4 |
| **MATSUDA** | female | 14.22 | 8.42 | 31.17 | 19.09 |
| **SECR_STUM** | female | 956.39 | 552.93 | 910.11 | 330.12 |
| **TRIG** | female | 1.23 | 0.67 | 1.15 | 0.6 |
| **WAIST** | female | 92.1 | 13.97 | 78.39 | 12.1 |
| **AUC_GLUCOSE** | male | 729.52 | 114.2 | 643.57 | 100.82 |
| **BMI** | male | 30.12 | 5.32 | 25.27 | 3.17 |
| **HDL** | male | 1.2 | 0.25 | 1.34 | 0.35 |
| **HIP** | male | 105.41 | 11.89 | 97.37 | 5.96 |
| **INSU_F** | male | 9.38 | 6.61 | 6.66 | 4.62 |
| **MATSUDA** | male | 13.53 | 9.04 | 32.69 | 21.9 |
| **SECR_STUM** | male | 917.41 | 574.45 | 849.22 | 339.83 |
| **TRIG** | male | 1.59 | 1.06 | 1.49 | 0.88 |
| **WAIST** | male | 102.84 | 13.56 | 89.97 | 9.37 |

TRIG: fasting triglycerides in mmol/l; HDL: HDL-cholesterol in mmol/l; INSU_F: fasting insulin in pmol/l; SECR_STUM: Stumvoll's first phase insulin secretion index computed as 2503+6.476 * INSU_F - 126.5 * GLUC_2h + 0.954 * INSU_2h - 293.3 * GLUC_F MATSUDA: insulin sensitivity computed as 10000/sqrt(GLUC_F*INSU_F*(INSU_F+INSU_2h)/2*(GLUC_F+GLUC_2h)/2); AUC_glucose: OGTT 5-point glucose (TUEF) or 2-point glucose area-under-curve computed according to the trapezoid rule;  BMI: body mass index in $kg/m^2$ ; HIP and WAIST circumferences in cm

#### Supplementary Table 15

Cluster medians (sex-stratified and Z-score-transformed value for each clustering variable on the population means/sd) in TUEF/TULIP (n=899) and in Whitehall-II (n=6810) for the proxy clustering variables

| | CLUSTER 1 TUEF | CLUSTER 1 WH-II | CLUSTER 2 TUEF | CLUSTER 2 WH-II | CLUSTER 3 TUEF | CLUSTER 3 WH-II | CLUSTER 4 TUEF | CLUSTER 4 WH-II | CLUSTER 5 TUEF | CLUSTER 5 WH-II | CLUSTER 6 TUEF | CLUSTER 6 WH-II |
|---|---|---|---|---|---|---|---|---|---|---|---|---|
| **AUC_GLU** | -0.435 | -0.575 | -0.59 | -1.076 | 0.699 | 1.02 | -0.667 | -1.021 | 1.25 | 0.967 | 0.143 | -0.451 |
| **BMI** | -0.475 | -0.761 | -1.196 | -1.149 | -0.244 | -0.504 | 0.165 | -0.027 | 0.692 | 0.192 | 0.777 | 0.373 |
| **HDL** | -0.14 | -0.355 | 1.08 | 1.034 | -0.101 | 0.046 | -0.396 | -0.395 | -0.762 | -0.877 | -0.323 | -0.315 |
| **HIP** | -0.455 | -0.666 | -1.016 | -0.93 | -0.161 | -0.463 | 0.344 | 0.016 | 0.597 | -0.144 | 0.693 | 0.195 |
| **INSU_F** | -0.369 | -0.163 | -0.806 | -0.814 | -0.282 | -0.245 | -0.438 | -0.375 | 0.667 | 1.152 | 0.58 | 1.134 |
| **MATSUDA** | -0.092 | 0.317 | 1 | 2.551 | -0.449 | -0.123 | 0.272 | 1.241 | -0.967 | -0.756 | -0.861 | -0.499 |
| **SECR_STUM** | -0.164 | 0.155 | -0.525 | -0.308 | -0.549 | -0.38 | -0.238 | -0.056 | 0.38 | 0.702 | 0.435 | 1.006 |
| **TRIG** | -0.278 | 0.184 | -0.699 | -0.553 | -0.129 | 0.15 | -0.406 | 0.005 | 0.489 | 1.542 | 0.071 | 0.33 |
| **WAIST** | -0.579 | -0.785 | -1.224 | -1.316 | -0.007 | -0.517 | 0.136 | 0.011 | 0.923 | 0.292 | 0.78 | 0.482 |

For variable description see previous Supplementary Table
